# A novel *UNC93B1* gain-of-function mutation leads to TLR7 and TLR8 hyperactivation and systemic lupus erythematosus

**DOI:** 10.1101/2024.09.11.24313360

**Authors:** Xu Han, Ruoran Wang, Seza Ozen, Qintao Wang, Wei Dong, Yi Zeng, Ouyuan Xu, Seher Şener, Li Guo, Ying Gu, Huanming Yang, Xiaomin Yu, Panfeng Tao, Qing Zhou

**Affiliations:** Liangzhu Laboratory, Zhejiang University, and Department of Rheumatology, The Second Affiliated Hospital, Zhejiang University School of Medicine, Hangzhou, China; Department of Pediatrics, Division of Pediatric Rheumatology, Hacettepe University Faculty of Medicine, Ankara, Turkey; Hangzhou Institute of Medicine Chinese Academy of Sciences, Hangzhou, China; BGI, Shenzhen, China; Life Sciences Institute, Zhejiang University, Hangzhou, China; Department of Rheumatology Immunology & Allergy, Children’s Hospital, National Clinical Research Center for Child Health, Zhejiang University School of Medicine, China

## Abstract

Dysfunctions in nucleic acid-sensing Toll-like receptors (TLRs) disrupt nucleic acid tolerance, leading to systemic lupus erythematosus (SLE). Here, we report a novel homozygous gain-of-function p.R95L mutation in the TLR chaperone protein UNC93B1 in an SLE patient. Bulk and single-cell transcriptional analysis of the patient’s peripheral blood mononuclear cells (PBMCs) revealed significantly elevated inflammation in T cells and myeloid cells due to enhanced dendritic cells function. The *UNC93B1* R95L mutation leads to hyperactivation of TLR7/8, but not TLR3/9, upon stimulation with specific agonists *in vitro*. Transgenic *Unc93b1^R95L/R95L^* mice develop inflammatory and autoimmune phenotypes, and the upregulation of inflammatory signaling differs among organs, with a specific contribution of malfunctioned T cells and B cells. In human and mouse cell lines, the *UNC93B1* R95L mutation promotes TLR7/8 signaling by enhancing its binding to ssRNA, without affecting TLR7/8 translocation. Overall, our results elucidate the pathology of organs and the immunological profiles of immune cell populations in the development of SLE caused by the *UNC93B1* R95L mutation through the TLR7/8 axis.

## Introduction

Systemic lupus erythematosus (SLE) is a chronic autoimmune disease characterized by autoantibodies targeting nuclear antigens and affecting multiple organ systems (1). Monogenic lupus refers to a group of diseases with lupus-like symptoms caused by mutations in single genes. These genes mainly affect complement pathways, type I interferon (IFN) signaling, nucleic acid sensing, RAS signaling pathways, and immune tolerance(2). Identifying these pathogenic genes is crucial for improving SLE classification and treatment. Genetic diagnosis enables personalized treatment, and understanding the mechanisms and effects of these mutations provides insights into SLE and other autoimmune diseases, aiding in the development of targeted therapies.

Toll-like receptors (TLRs) are critical for recognizing microbial molecules and initiating immune responses. They consist of an extracellular domain for ligand recognition, a transmembrane domain, and a Toll-interleukin 1 receptor (TIR) domain for signaling. Upon ligand binding, the TIR domain dimerizes and recruits proteins to form the Myddosome complex, which activates pathways such as NF-κB and type I IFN(3).

A subset of TLRs recognizes various forms of nucleic acids: TLR3 binds double-stranded RNA, TLR7 and TLR8 bind single-stranded RNA (ssRNA) fragments, and TLR9 binds CpG single-stranded DNA(4). TLR7, found in dendritic cells (DCs), monocytes, and B cells, recognizes ssRNA within endosomes(5, 6). Loss-of-function (LOF) mutations in *TLR7* cause immunodeficiency(7), while gain-of-function (GOF) mutations lead to abnormal activation of NF-κB and type I IFN pathways, resulting in SLE-like symptoms such as autoimmune thrombocytopenia, elevated autoantibodies, complement deficiencies, arthritis, migraines, and kidney involvement(8–10). TLR8, expressed in myeloid DCs, monocytes, and neutrophils(11), also recognizes ssRNA in endosomes. GOF mutations in TLR8 cause immunodeficiency and inflammation(12). Patients show immunodeficiency symptoms like abnormal B cell differentiation, neutropenia, and infections, alongside inflammatory symptoms like arthritis, vasculitis, fever, T cell activation, increased cytokine production, and elevated NF-κB signaling. Tlr8 in mice is generally considered non-functional, and little is known about its regulation and role in autoimmune diseases.

UNC93B1 is a chaperone protein crucial for transporting TLR3, TLR7, TLR8, and TLR9 from the endoplasmic reticulum (ER) to endosomes where these receptors initiate signaling(13, 14). Mice with homozygous *Unc93b1^H412R^* mutation are more susceptible to pathogens due to impaired binding of Unc93b1 to TLR3, TLR7, and TLR9, trapping them in the ER and blocking signal transduction(15). Patients with *UNC93B1* LOF mutations are prone to herpes simplex encephalitis due to dysfunctional endosomal TLRs, with reduced responses of immune cells to TLR agonists(16). Understanding the role of UNC93B1 in TLR regulation is critical for elucidating its involvement in autoimmune diseases like SLE. Recent structures have identified potential key regions involved in UNC93B1-TLR7 interactions(17), but the significance of these residues in UNC93B1 and TLR7 function remains unexplored.

Here, we identify a patient with early-onset SLE carrying homozygous *UNC93B1* R95L mutation. Through a series of experiments in human and mouse model, we elucidate the pathology of organs and the immunological profiles of immune cell populations in the development of SLE caused by the *UNC93B1* R95L mutation through the TLR7/8 axis.

## Results

### A novel *UNC93B1* pathogenic mutation in a patient with early-onset SLE

The proband is a female who presented with symptoms suggesting early-onset SLE. These symptoms included fevers, fatigue, discoid and maculopapular rashes, and photosensitivity (**Figure 1A**). Immunologic studies revealed positive anti-nuclear antibody (ANA), anti-double-stranded DNA (anti-dsDNA) antibodies exceeding 800 U/L, as well as positive anti-Smith (anti-Sm) and anti-Ribonucleoprotein (anti-RNP) antibodies. The patient had hypocomplementemia with reduced levels of C3 (0.33 g/L, normal range 0.8-1.6 g/L) and C4 (0.05 g/L, normal range 0.15-0.4 g/L). C-reactive protein (CRP) level was normal, her erythrocyte sedimentation rate (ESR) was elevated, ranging between 40-60 mm/hour (normal range 1-10 mm/hour). Based on the 2019 EULAR/ACR criteria(18), she was diagnosed with early-onset SLE. Currently she is on prednisolone at a dosage of 5 mg/day, hydroxychloroquine at 150 mg/day, and mycophenolate mofetil at 750 mg/day.

**Figure 1.**
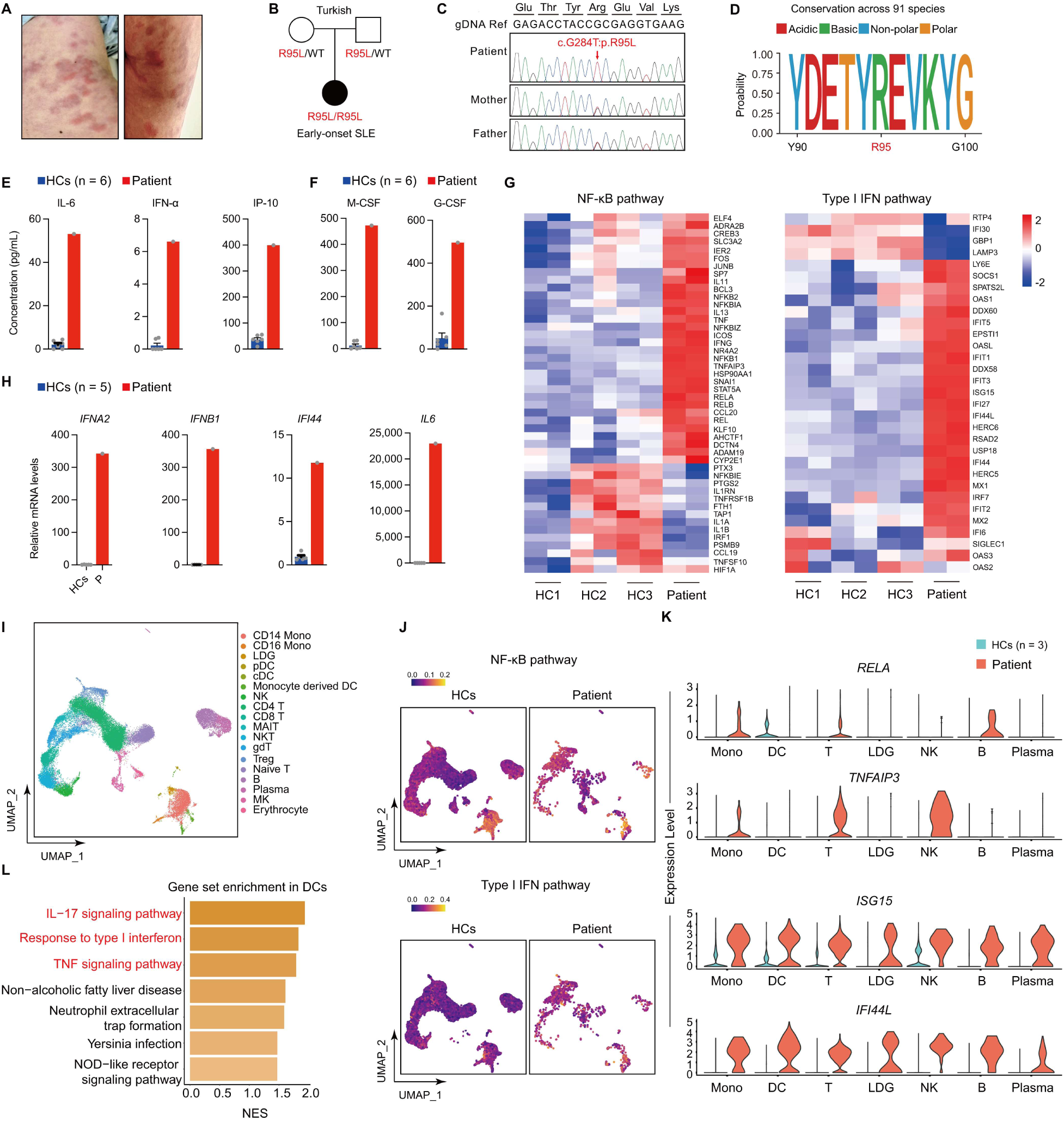
Inflammatory signaling activated in an early-onset SLE patient with *UNC93B1* R95L mutation. (A) Clinical images displaying skin rashes on the patient’s back (left) and hip (right). (B) Pedigree of the patient with a homozygous variant c.284G>T (NM_030930), p.R95L in *UNC93B1*. (C) Validation of the homozygous variant in *UNC93B1* using Sanger sequencing. (D) The evolutionary conservation of the arginine at position 95 in UNC93B1 across species. (E) Serum Levels of cytokines IL-6, IFN-α, and chemokine IP-10 in the patient (P) and healthy controls (HCs, n = 6) were detected by CBA. (F) Serum Levels of colony-stimulating factors M-CSF and G-CSF in the patient (P) and healthy controls (HCs, n = 6) were detected by ELISA. (G) Transcription level of NF-κB and type I IFN pathways in PBMCs from the patient (P) and healthy controls (HC, n = 3). Analysis of each sample was performed in duplicate. (H) qPCR analysis of NF-κB and type I IFN pathways related genes in PBMCs from the patient (P) compared with healthy controls (HCs, n = 5). (I) Uniform manifold approximation and projection (UMAP) visualization and marker-based annotation of 18 cell subtypes from the patient and healthy controls (HCs, n = 3). (J) UMAP visualization of the target pathway scoring in the patient and healthy controls (HCs, n = 3). Upper, pathway scoring based on genes on NF-κB signal transduction gene set in Gene Ontology database; bottom, pathway scoring based on genes on response to type I IFN gene set in Gene Ontology database. Scores were generated based on Seurat Addmodulescore method. (K) Expression levels of essential genes in NF-κB and type I IFN pathways in various immune cells among the patient and healthy controls (HCs, n = 3). (L) Enrichment of upregulated pathways of differential expressed gene in patient’s DCs based on GSEA.

Whole-exome sequencing of the proband and her parents identified a homozygous mutation in *UNC93B1*: NM_030930:c.284G>T, p.Arg95Leu (R95L) (**Figure 1, B and C**). Her healthy parents were heterozygous carriers of this variant. The R95 residue is highly conserved across species (**Figure 1D**). This mutation was not found as homozygous in public databases, including gnomAD v4, Kaviar, and chinaMap. *In silico* modeling predicted that this mutation impacts protein function, with a CADD score of 35 and a GERP score of 4.55, both indicating a deleterious effect.

### Activation of inflammatory signaling in the patient

We obtained serum samples from the patient and quantified cytokines using cytometric bead array (CBA) and enzyme-linked immunosorbent assay (ELISA). The analysis revealed markedly increased production of proinflammatory cytokines IL-6 and IFN-α, chemokine IP-10, and macrophage colony-stimulating factors (M-CSF) and Granulocyte-CSF (G-CSF) compared to healthy controls (**Figure 1, E and F**).

To examine the transcriptional differences between the patient and healthy controls, we performed bulk RNA sequencing on peripheral blood mononuclear cells (PBMCs) samples. Differential gene expression analysis showed that expression of inflammatory response genes was elevated in patient compared to healthy controls (**Supplemental Figure 1A**). Gene set enrichment analysis (GSEA) showed that type I IFN and NF-κB signaling were top-ranked among the upregulated pathways in the patient (**Supplemental Figure 1B**). The heightened expression of genes in type I IFN and NF-κB pathways were also observed in patient (**Figure 1G, Supplemental Figure 1A)**. These results suggested specific effect of these two pathways in the patient’s inflammatory manifestation. The changes in the type I IFN pathway genes were notably pronounced. Quantitative PCR (qPCR) confirmed increased transcription of type I IFN, including *IFNA2* and *IFNB1*, type I IFN-stimulated gene (ISG) *IFI44*, as well as *IL6*, which is regulated by the NF-κB pathway, in the patient compared to healthy controls (**Figure 1H**). Many SLE patients exhibit elevated levels of type I IFN, which correlates with the secretion of autoantibodies, disease severity, and heterogeneity(19). Thus, the abnormal activation of the type I IFN signaling pathway may be a critical factor contributing to the patient’s SLE.

To further identify specific cell populations and immunological pathways involved in the patient’s pathology, we analyzed the single-cell transcriptomes of PBMCs from the patient and three healthy controls. Based on the integration and unsupervised clustering of single-cell sequencing data, we identified 18 cell populations according to specific profiles of marker gene expression (**Figure 1I, Supplementary** Figure 1C). Compared to healthy controls, patient’s PBMCs showed elevated signal in NF-κB and type I IFN pathways (**Figure 1J**). Differential expression analysis displayed a distinct expression pattern of key genes in NF-κB pathway, such as *RELA*, *TNFAIP3* in myeloid cells and T cells, and critical genes in type I IFN pathway, including *ISG15*, *IFI44L* in most immune cells (**Figure 1K)**. Among myeloid cells, plasmacytoid DCs (pDCs) are the primary interferon-producing cells(20), whereas monocyte-derived DCs (moDCs) and conventional DCs (mDCs) could activate T cells through antigen presentation(21, 22). Enrichment of IL-17 and type I IFN signaling in DCs and T cells implicated specific contributions of DCs subsets, pDCs, mDCs, and moDCs, to the patient’s manifestation through T cell activation and type I IFN production (**Figure 1L, Supplemental Figure 1D**).

These results demonstrated dysregulated inflammatory regulation in the patient, suggesting that *UNC93B1* R95L mutation plays a driving role in the activation of immune cell responses.

### *UNC93B1* R95L mutation promotes inflammation through TLR7/TLR8 activation

Unrestrained intracellular TLR activation can lead to responses to self-nucleic acids and autoimmunity(8). To investigate the effect of the *UNC93B1* R95L mutation on intracellular TLRs (TLR3, TLR7, TLR8, TLR9), we co-expressed wild-type UNC93B1, UNC93B1 R95L mutant, and UNC93B1 H412R mutant (negative control) with different TLRs in HEK293T cells. We measured the NF-κB reporter activity and/or the levels of IL-8 transcription following stimulation with specific TLR agonists (TLR3: Poly(I:C); TLR7/TLR8: R848; TLR9: ODN2216). These assays revealed that the *UNC93B1* R95L mutation enhanced the activation of TLR7, TLR8, and TLR9, but not TLR3 (**Figure 2, A-D**).

**Figure 2.**
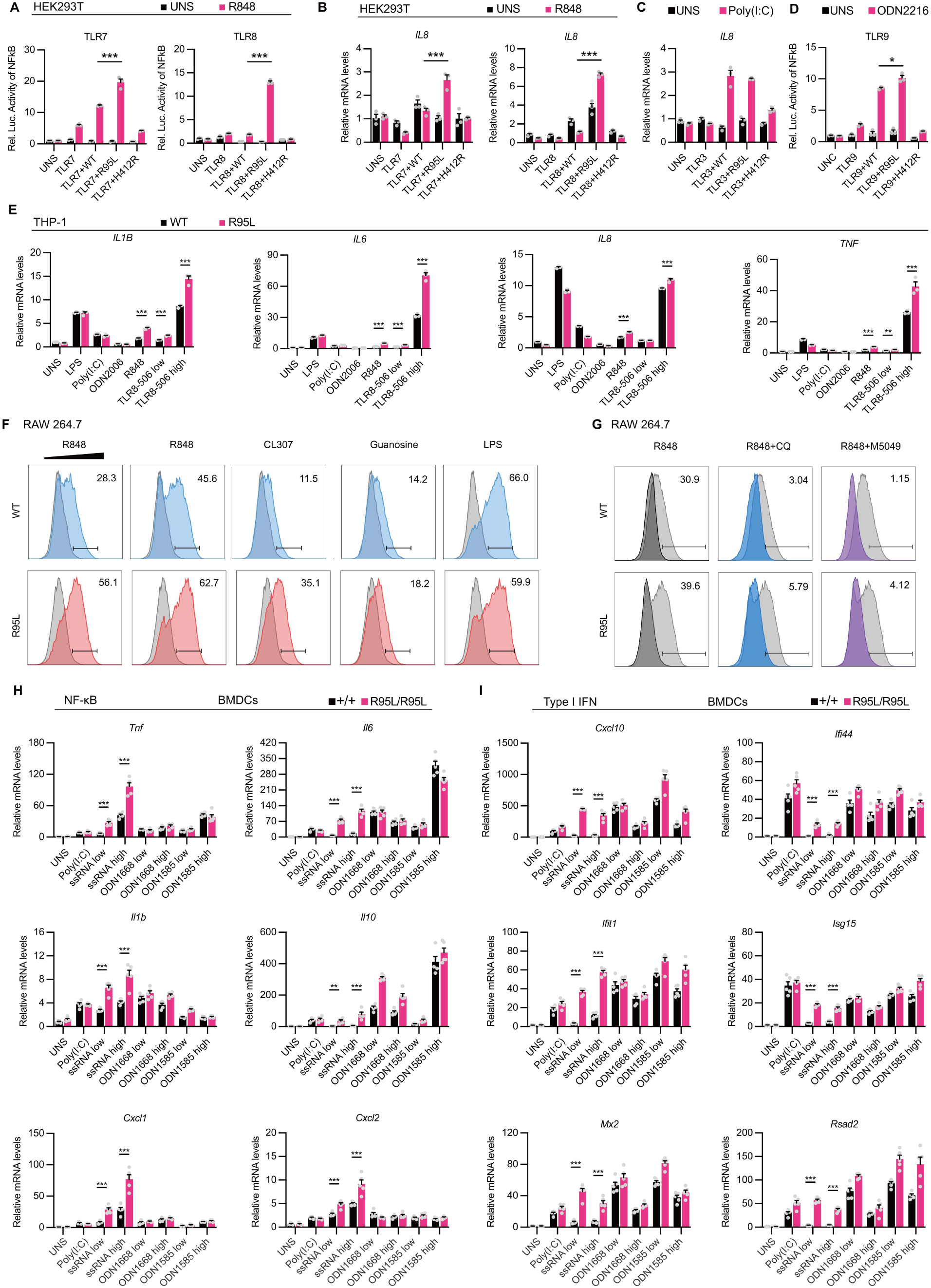
*UNC93B1^R95L^* mutation promotes inflammation through TLR7/TLR8 activation. (A) NF-κB luciferase assays in HEK293T cells co-transfected TLR7 (left) or TLR8 (right) with different UNC93B1 expression plasmids, treated as indicated (n = 3 biological replicates). Data represent mean ± SEM; **, *P* < 0.01; ***, *P* < 0.001; two-tailed unpaired Student’s *t* test. EV: empty vector; WT: wild-type UNC93B1; R95L: R95L mutant; HR: H412R mutant. (B) qPCR analysis of *IL8* transcription level in HEK293T cells co-transfected TLR7 (left) or TLR8 (right) with different UNC93B1 expression plasmids, treated as indicated (n = 3 biological replicates). Data represent mean ± SEM; ***, *P* < 0.001; two-tailed unpaired Student’s *t* test. (C) qPCR analysis of *IL8* in HEK293T cells co-transfected TLR3 with different UNC93B1 expression plasmids, treated as indicated (n = 3 biological replicates). Data represent mean ± SEM; two-tailed unpaired Student’s *t* test. EV: empty vector; WT: wild-type UNC93B1; R95L: R95L mutant; HR: H412R mutant. (D) NF-κB luciferase assay in HEK293T cells co-transfected TLR9 with different UNC93B1 expression plasmids, treated as indicated (n = 3 biological replicates). Data represent mean ± SEM; *, *P* < 0.05; two-tailed unpaired Student’s *t* test. WT: wild-type UNC93B1; R95L: R95L mutant. (E) qPCR analysis of *IL1B*, *IL6*, *IL8* and *TNF* transcription levels in THP-1 expressing the WT or R95L mutant UNC93B1 treated with various TLRs agonists (n = 3 biological replicates). Data represent mean ± SEM; **, *P* < 0.01; ***, *P* < 0.001; two-tailed unpaired Student’s *t* test. (F, G) Intracellular cytokine staining of Tnf in mouse RAW 264.7 expressing the WT or R95L mutant UNC93B1, and treated with various TLRs agonists or inhibitors. (H, I) qPCR analysis of NF-κB (H) and type I IFN (I) pathways related genes transcription in BMDCs from indicated mice treated with various TLRs agonists(n = 5 mice per group). Data represent mean ± SEM; **, *P* < 0.01; ***, *P* < 0.001; two-tailed unpaired Student’s *t* test.

In the human monocytic cell line THP-1, the UNC93B1 R95L mutant enhanced activation by the TLR7/8 dual agonist R848 and the TLR8-specific agonist TLR8-506, while there was no significant effect on TLR3 and TLR4, which are UNC93B1-independent (**Figure 2E**). In mouse macrophage cell line RAW 264.7 (where TLR8 is non-functional), the UNC93B1 R95L mutant increased TLR7 activation upon R848 stimulation, significantly elevating the transcription levels of NF-κB downstream genes such as *Tnf* and *Il1b* (**Supplemental Figure 2A**). Intracellular cytokine staining of Tnf further confirmed that the UNC93B1 R95L mutant enhanced TLR7 activation by various TLR7 agonists (**Figure 2F**). Both chloroquine, which inhibits TLR activation by altering endosomal pH and inhibiting the ectodomain cleavage, and M5049, a small molecule inhibitor of TLR7/8, effectively suppressed R95L-induced TLR7 activation and its downstream signaling (**Figure 2G**).

Bone marrow-derived dendritic cells (BMDCs) and macrophages (BMDMs) from the homozygous *Unc93b1^R95L^* mice showed enhanced activation of NF-κB and type I IFN signalings in response to TLR7 ligands but not ligands of TLR3, TLR4, and TLR9 (**Figure 2, H and I, Supplemental Figure 2B**). Interestingly, in BMDCs, the transcription levels of both NF-κB and type I IFN-related genes were significantly upregulated, whereas in BMDMs, type I IFN-related gene expression showed no significant changes.

Collectively, experiments across human cell lines, mouse cell lines, and primary isolated mouse cells demonstrate that the *UNC93B1* R95L mutation confers a gain-of-function effect with selectively enhanced TLR7/8 but not TLR3/9 activation, leading to downstream NF-κB and type I IFN signaling dysregulation.

### Mice with *Unc93b1^R95L^* mutation develop systemic inflammation and autoimmune pathology

To verify that the *UNC93B1* R95L mutation drives autoimmune manifestation and abnormal inflammatory signaling, we generated *Unc93b1^R95L/R95L^*knock-in mice on a C57BL/6J background using CRISPR/Cas9 technology (**Figure 3A**). We monitored mice body weight and cutaneous manifestation weekly, and measured plasma anti-dsDNA antibody level biweekly from postnatal day 21 until the phenotype stabilized. Over a 9-week period, no significant difference in body weight was observed between mutant and control mice, regardless of sex (**Figure 3B**). Tissue analysis revealed that *Unc93b1^R95L/R95L^*mice exhibited significant splenomegaly and slight hepatomegaly compared to controls, whereas *Unc93b1^+/R95L^* mice showed no differences (**Figure 3C, Supplemental Figure 3A**). Elevated levels of autoantibodies (including dsDNA, dsRNA, and ANA), C-reactive protein, and C3 were observed in *Unc93b1^R95L/R95L^* mice compared to *Unc93b1^+/+^*controls, as measured by ELISA (**Figure 3D**). Given the weaker phenotype observed in males, we focused subsequent inflammation analysis on female mice. Plasma cytokines and chemokines analysis revealed elevated levels of pro-inflammatory cytokines Il-6 and Il-1β, as well as the chemokine Cxcl1, in *Unc93b1^R95L/R95L^* mice compared to *Unc93b1^+/+^* controls (**Figure 3E**). These data indicate that the *Unc93b1^R95L^* mutation leads to autoimmune and inflammatory phenotypes in mice, recapitulating the clinical symptoms of the patient.

**Figure 3.**
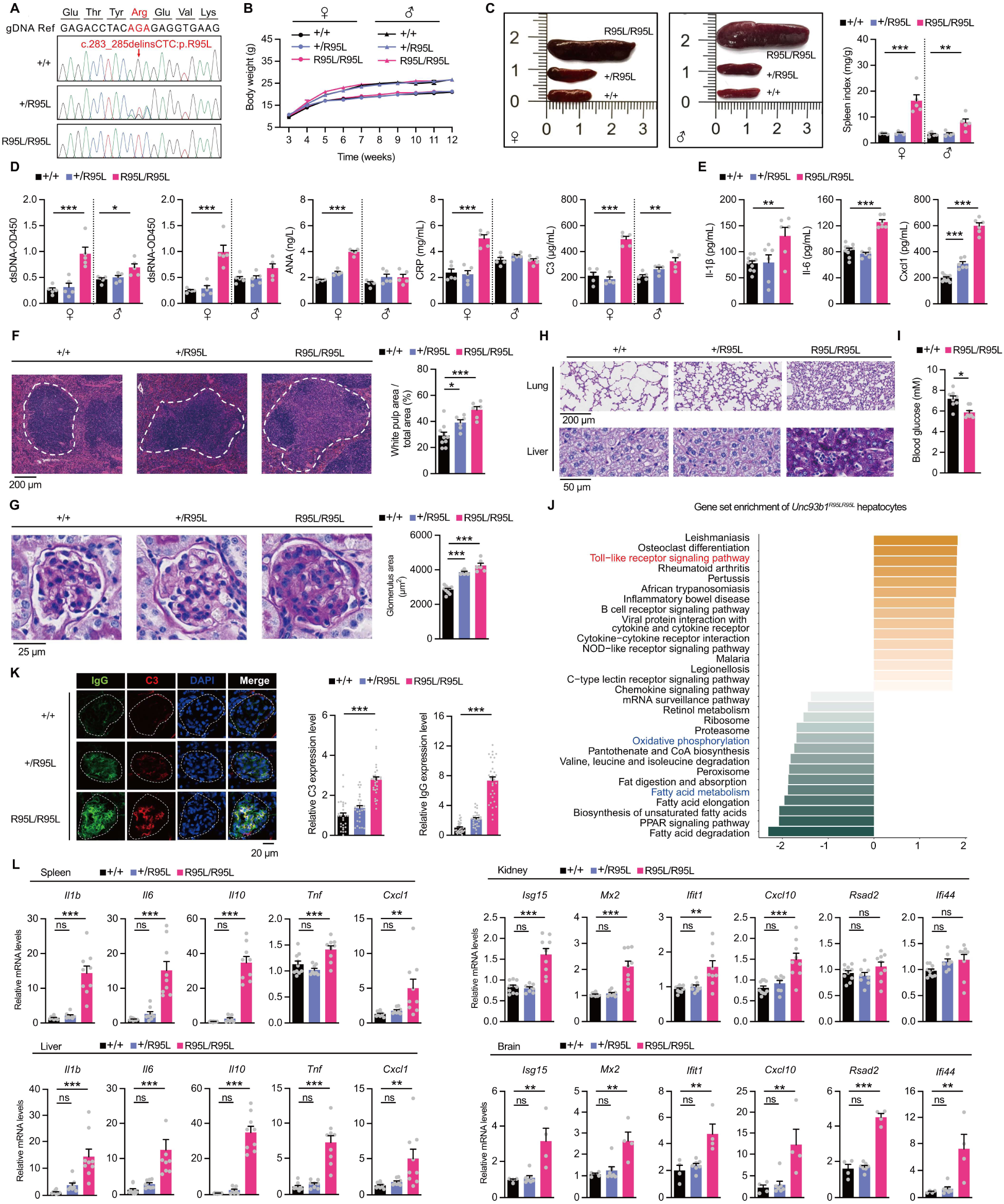
Mice with *Unc93b1^R95L^*mutation develop systemic inflammation and autoimmune pathology. (A) Sanger sequencing showing the amino acid substitution *Unc93b1,* ENSMUST00000162708.7: c.283_285delinsCTC, p.R95L in homozygous knock-in mice. Wild type (+/+), heterozygous (+/R95L), and homozygous (R95L/R95L) are indicated. (B) Body weight of *Unc93b1^+/+^, Unc93b1^+/R95L^* and *Unc93b1^R95L/R95L^* mice. (C) Representative images of splenomegaly and normalized spleen weight to body weight (spleen index) of 12-wk-old mice (n = 5 per group). Data represent mean ± SEM; **, *P* < 0.01; ***, *P* < 0.001; one-way ANOVA. (D) Plasma levels of anti-DNA, anti-RNA, ANA, CRP and C3 in the indicated mice were detected using ELISA (n = 5 per group). Data represent mean ± SEM; *, *P* < 0.05; **, *P* < 0.01; ***, *P* < 0.001; one-way ANOVA. (E) Plasma levels of Il-1β, Il-6 and chemokine Cxcl1 in the indicated mice were detected using ELISA (*Unc93b1^+/+^,* n = 10; *Unc93b1^+/R95L^*, n = 6; *Unc93b1^R95L/R95L^*, n = 6). Data represent mean ± SEM; **, *P* < 0.01; ***, *P* < 0.001; one-way ANOVA. (F) H&E staining of the spleen from indicated mice (left). Normalized white pulp was calculated (right). (*Unc93b1^+/+^*, n = 10; *Unc93b1^+/R95L^*, n = 6; *Unc93b1^R95L/R95L^*, n = 6). Data represent mean ± SEM; *, *P* < 0.05; ***, *P* < 0.001; one-way ANOVA. (G) PAS staining of the kidney from indicated mice (left). Glomerulus area was calculated (right). (*Unc93b1^+/+^*, n = 10; *Unc93b1^+/R95L^*, n = 6; *Unc93b1^R95L/R95L^*, n = 6).Data represent mean ± SEM; ***, *P* < 0.001; one-way ANOVA. (H) PAS staining of the lung and liver from indicated mice. (I) Short fasting blood glucose levels of the indicated mice (n = 8 per group). Data represent mean ± SEM; **, *P* < 0.01; two-tailed unpaired Student’s *t* test. (J) Top 15 enriched pathways of differential expressed genes in murine hepatocytes from the *Unc93b1^+/+^*, and *Unc93b1^R95L/R95L^* mice. (K) Representative immunofluorescent images of IgG and C3 in glomerulus from the indicated mice(left). IgG (top) and C3 (bottom) staining in individual glomerulus was quantified (n = 30 glomerulus from three mice per group). Data represent mean ± SEM; ***, *P* < 0.001; one-way ANOVA. (L) qPCR analysis of NF-κB and type I IFN pathways related genes in the spleen, kidney, liver, and brain from the indicated mice (spleen, kidney and liver, n = 8-10 per group; brain,n = 4-7 per group). Data represent mean ± SEM; **, *P* < 0.01; ***, *P* < 0.001; one-way ANOVA.

To further investigate the pathological mechanisms underlying the autoimmune and inflammatory symptoms in *Unc93b1^R95L/R95L^* mice, we analyzed the pathological differences in the spleen, kidney, lung, liver, brain, quadriceps, and plasma, comparing *Unc93b1^R95L/R95L^* mice with littermate controls.

Histological examination using hematoxylin and eosin (H&E) staining revealed significantly increased number of splenocytes and enlarged white pulp areas in the spleens of both *Unc93b1^+/R95L^* and *Unc93b1^R95L/R95L^*mice, with indistinct boundaries between the white and red pulp (**Figure 3F**). Periodic acid-Schiff (PAS) staining of kidneys showed significantly increased glomerulus areas in both *Unc93b1^+/R95L^* and *Unc93b1^R95L/R95L^*mice, with inflammatory cell infiltration in the renal interstitial area, particularly around renal tubules in *Unc93b1^R95L/R95L^* mice (**Figure 3G)**. This was accompanied by glomerular damage characterized by increased cell numbers, immune complex deposition, and expansion of mesangial matrix (**Supplemental Figure 3B**). Lung tissue analysis showed reduced alveolar spaces and thickened septa in both *Unc93b1^+/R95L^* and *Unc93b1^R95L/R95L^* mice, with pulmonary congestion noted in *Unc93b1^R95L/R95L^*mice. The liver of *Unc93b1^R95L/R95L^* mice exhibited prominent glycogen storage within hepatocytes and marked inflammatory cell infiltration (**Figure 3H**). These observations were consistent with significant hypoglycemia in *Unc93b1^R95L/R95L^*mice (**Figure 3I**). GSEA further revealed an upregulation of inflammatory signaling and downregulation of glycogen metabolism downstream signaling in *Unc93b1^R95L/R95L^*mice liver (**Figure 3J**). Immunofluorescence staining of glomeruli revealed elevated IgG and C3 deposition in *Unc93b1^R95L/R95L^* mice, while *Unc93b1^+/R95L^* mice showed a slight increase in IgG deposition (**Figure 3K**). These findings, aligned with PAS staining results, confirm immune complex deposition and highlight the autoimmune pathology in *Unc93b1^R95L/R95L^* mice.

These analyses suggest that the *Unc93b1^R95L^* mutation leads to widespread tissue inflammation and immune cell infiltration in multiple organs.

### Differential activation of NF-κB and type I IFN pathways in various tissues

In the patient’s PBMCs, we observed a more ubiquitous upregulation of genes in type I IFN pathway compared to NF-κB pathway, implying differential activation patterns of these pathways through the dysfunctional UNC93B1/TLR axis in various tissues. To investigate the effects of the *UNC93B1* R95L mutation on inflammatory pathways in different tissues, we performed qPCR to measure the transcription levels of downstream genes in various tissues of the mice.

Interestingly, the *Unc93b1^R95L^*mutation significantly enhanced the activation of NF-κB pathway in the spleen and liver (**Figure 3L**, **Supplemental Figure 3C**). Additionally, upregulated genes in splenocytes from *Unc93b1^R95L/R95L^*mice exhibited enrichment in inflammatory and autoimmune signaling pathways, implicating a specific contribution of splenocytes to immune dysregulation (**Supplemental Figure 3D**). In contrast, the mutation significantly enhanced the activation of the type I IFN pathway in the kidney and brain (**Figure 3L**). Despite the morphological evidence of substantial lung tissue damage, qPCR results indicated that both the NF-κB and type I IFN pathways were less pronounced in the lungs compared to other tissues. The quadriceps muscle served as a negative control with no differential expression of genes related to either pathway (**Supplemental Figure 3E**).

These findings suggest that the *Unc93b1^R95L^* mutation differentially affects NF-κB and type I IFN signaling pathways across various tissues, contributing to the complex autoimmune and inflammatory phenotype observed in these mice.

### Effects of *Unc93b1^R95L^* mutation on immune cell populations in mice

Since the *Unc93b1^R95L^* mutation affected inflammatory signaling in various tissues, we next explored the impact of *Unc93b1^R95L^*mutation on exact immune cell populations to elaborate on its pathogenesis to SLE. We performed single-cell sequencing on murine spleen tissue from *Unc93b1^R95L/R95L^*and *Unc93b1^+/+^* mice. Unsupervised clustering identified 28 cell populations from murine splenocytes (**Figure 4A, Supplemental Figure 4A**). In view of globally activated inflammatory response and autoimmune manifestation in the spleen of *Unc93b1^R95L/R95L^* mice, we carried out differential expression analysis among cell populations between *Unc93b1^R95L/R95L^* and *Unc93b1^+/+^* mice. Amplified signaling in NF-κB pathway was observed in myeloid cells and T cells from *Unc93b1^R95L/R95L^* mice, with significant activation of NF-κB through pathways like TNF signaling, which suggests that *Unc93b1^R95L^* mutation drives systemic inflammation state through T cells (**Figure 4B**). Additionally, we also detected escalated production of immunoglobulin in B cells and plasma cells in *Unc93b1^R95L/R95L^* mice, with ascendant gene expression in the gene set about SLE pathogenesis, which implicated a promoting role of the *Unc93b1^R95L^* mutation in autoimmune pathology through B cells (**Figure 4C**). These results implicated a dysfunctinoal state of lymphocytes in *Unc93b1^R95L/R95L^*mice, which may contribute to the SLE pathogenesis.

**Figure 4.**
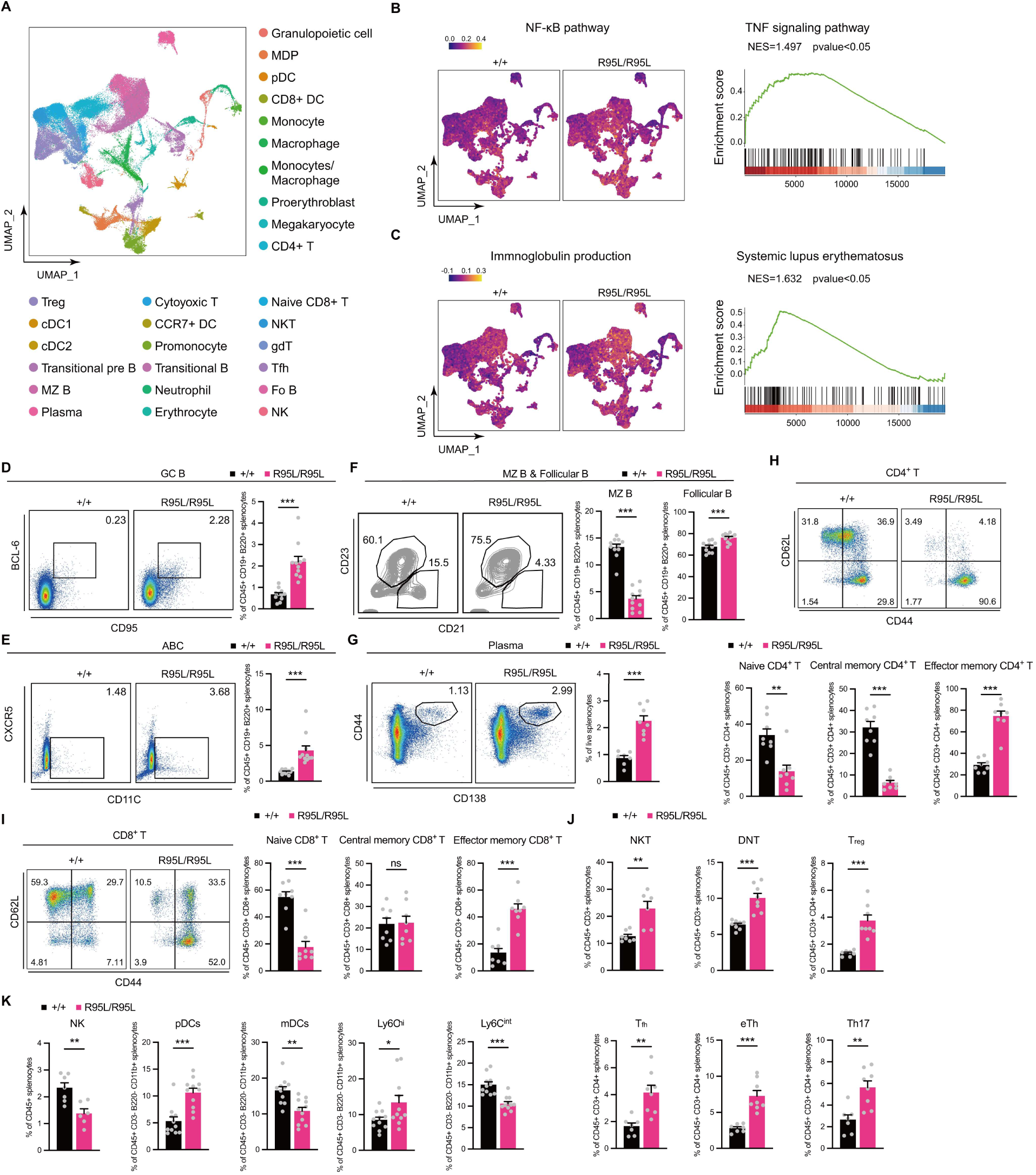
Effects of *UNC93B1^R95L^* mutation on immune cell populations in mice. (A) UMAP visualization and marker-based annotation of 28 cell subtypes from splenocytes of *Unc93b1^R95L/R95L^* and *Unc93b1^+/+^* mice (n = 3 mice per group). NKT, natural killer T cell; gdT, gamma-delta T cell; Treg, regulatory T cell; MZ B, marginal zone B cell; Fo B, follicular B cell; DC, dendritic cell; MDP, myeloid dendritic progenitor. (B) Upregulated inflammatory signal in *Unc93b1^R95L/R95L^* mice. Left, pathway scoring of NF-κB signal transduction gene set in Gene Ontology database; right, GSEA plot shows upregulated TNF signaling pathway in murine splenic T cells. Pathway score is calculated by the Seurat Addmodulescore method. (C) Upregulated autoimmune characteristics in *Unc93b1^R95L/R95L^* mice. Left, pathway scoring of immunoglobulin production gene set in Gene Ontology database; right, GSEA plot shows upregulated SLE signaling pathway in murine splenic B cells. Pathway score is calculated by the Seurat Addmodulescore method. (D-G) Splenic germinal center B cells (GC B) (D), age-associated B cells (ABC) (E), marginal zone B cells (MZ B) (F), follicular B cells (F), and plasma cells (G) distribution are indicated by FACS plots and percentages (n = 11 mice per group). Data represent mean ± SEM; ***, *P* < 0.001; two-tailed unpaired Student’s *t* test. (H, I) Splenic CD4^+^ T cells (H) and CD8^+^ T cells (I) activation are indicated by FACS plots and percentages (n = 8 mice per group). Data represent mean ± SEM; **, *P* < 0.01; ***, *P* < 0.001; two-tailed unpaired Student’s *t* test. (J) Flow cytometric analysis of splenic T cell populations from indicated mice (n = 6-8 mice per group). Data represent mean ± SEM; **, *P* < 0.01; ***, *P* < 0.001; two-tailed unpaired Student’s *t* test. (K) Flow cytometric analysis of splenic NK, pDCs, mDCs and monocytes populations from indicated mice (NK, n = 6-7 mice per group; pDCs, mDCs and monocytes, n = 11 mice per group). Data represent mean ± SEM; *, *P* < 0.05; **, *P* < 0.01; ***, *P* < 0.001; two-tailed unpaired Student’s *t* test.

To further investigate specific alterations of immune cells caused by the *Unc93b1^R95L^* mutation, we analyzed the immune phenotype in the spleen and peripheral blood of *Unc93b1^+/+^* and *Unc93b1^R95L/R95L^* mice. Single-cell analysis of splenocytes revealed an increase of Treg cells, follicular helper T (Tfh) cells, and plasma cells and a reduction of CD4^+^ T cells, naive CD8^+^ T cells in the *Unc93b1^R95L/R95L^*mice (**Supplemental Figure 4B**). Flow cytometry further confirmed a disruption in lymphocyte composition, with an increase in germinal center (GC) B cells, age-associated B cells (ABCs), follicular B cells, and plasma cells, indicating continuous activation of B cells in the spleens of *Unc93b1^R95L/R95L^* mice (**Figure 4, D-G, Supplemental Figure 5, A-C**), consistent with the high levels of autoantibodies found in the plasma of these mice.

The number of Naive T cells were reduced, whereas effector memory T cells have increased, suggesting a shift towards an activated or memory state, indicative of ongoing immune activation and chronic inflammation characteristics of SLE (**Figure 4, H and I, Supplemental Figure 5, D and E)**. Additionally, expansions in NKT cells, double-negative T (DNT) cells, Treg cells, effector Th (eTh) cells, and Th17 cells indicated the presence of inflammation and autoimmune responses in *Unc93b1^R95L/R95L^*mice. The rise in follicular helper T (Tfh) cells aligns with the expansion in GC B cells, reflecting an active autoimmune response (**Figure 4J, Supplemental Figure 5F**).

A reduction in NK cells, a cell type often impaired in SLE, suggests a dysfunction in immune regulation. An increase in pDCs number can lead to excess production of interferon, promoting inflammation and autoimmune responses in SLE. The decrease in mDCs, critical for antigen presentation and T cell activation, could be linked to abnormal antigen presentation dysfunctions and immune dysregulation. An expansion in Ly6C-high monocytes is associated with inflammation and tissue damage responses (**Figure 4K, Supplemental Figure 5G**). We also observed a further concentrated MHC-II signaling and upregulation of ICAM signaling from DCs to T cells (**Supplemental Figure 5, H and I**) in *Unc93b1^R95L/R95L^* mice splenocytes in single-cell analysis.

Collectively, these findings illustrate the altered immune landscape caused by the *UNC93B1* R95L mutation, highlighting its role in promoting systemic inflammation and autoimmunity.

### UNC93B1 R95L mutation enhances ligand binding to TLR7 by weakening the interaction between UNC93B1 and TLR7

We next examined how the UNC93B1 R95L mutation affects TLR7 and TLR8 activation. The structure of the TLR7-UNC93B1 complex reveals that the UNC93B1 R95L mutation is located within the H1 helix (residues 91-97) between the TM1 and TM2 transmembrane domains of UNC93B1 (**Figure 5A**). This region directly interacts with two loop regions of the C-terminal LRR-CT domain of TLR7 and exhibits strong shape complementarity(17). In HEK293T cells, immunoprecipitation and western blotting results indicated that the UNC93B1 R95L mutation significantly weakened the interaction between UNC93B1 and both TLR7 and TLR8 (**Figure 5, B and C**). We further investigated the interactions between UNC93B1 and endogenous TLR7. We generated RAW 264.7 cell lines stably expressing UNC93B1 (WT)-BirA* and UNC93B1 (R95L)-BirA* and performed streptavidin pull-down. Western blotting results showed that the UNC93B1 R95L mutation reduced the association between UNC93B1 and cleaved forms of TLR7 (**Figure 5D**).

**Figure 5.**
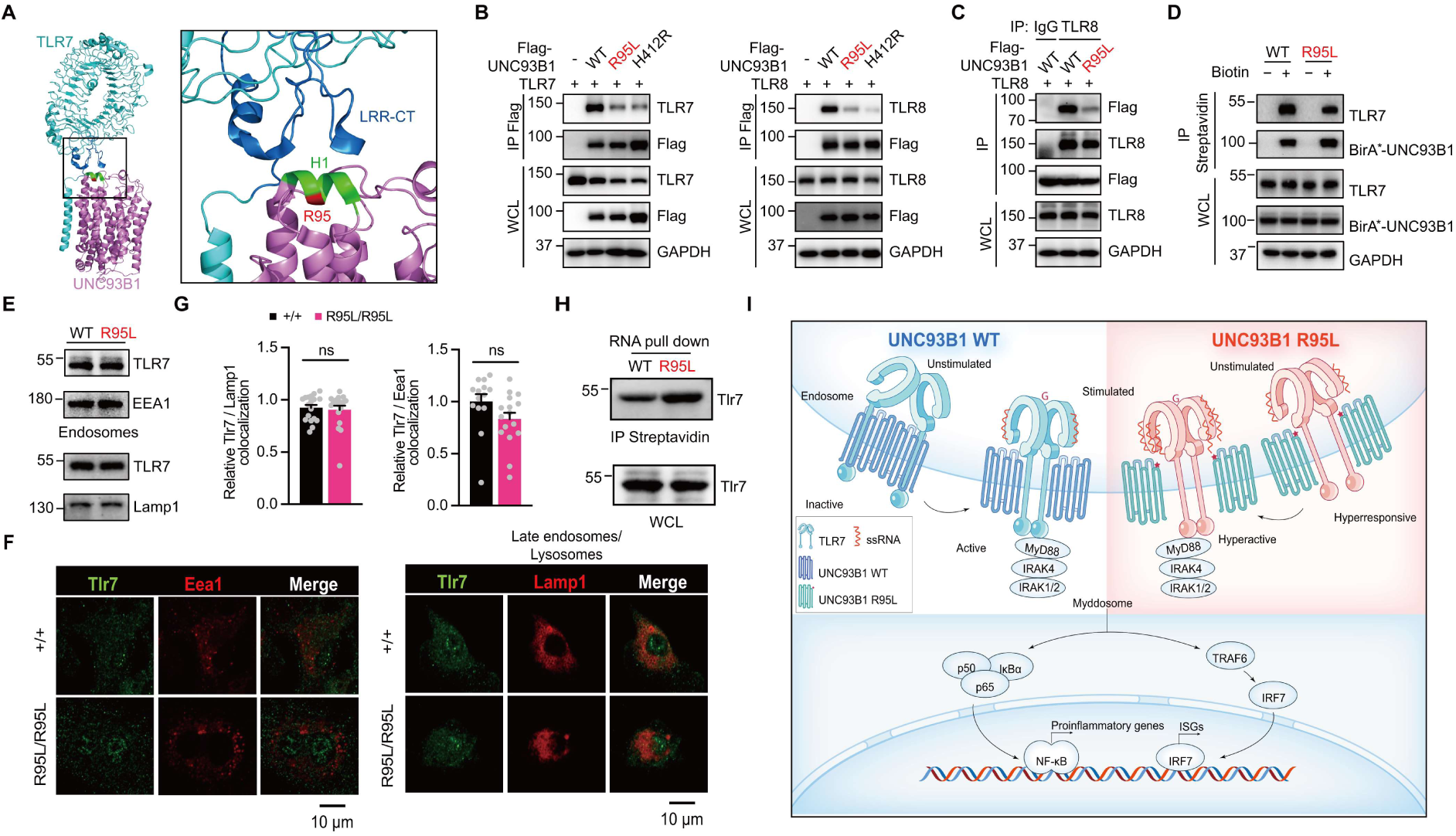
UNC93B1 R95L mutation enhances TLR7 ligand binding by weakening their interaction. (A) The structure of TLR7 and UNC93B1 complex (PDB: 7CYN) shows H1 helix (residues 91-97) of UNC93B1 directly contacting two loop regions of the C-terminal LRR-CT domain of TLR7. Protein structural analysis was executed with Pymol. (B) Flag-immunoprecipitation and western blot analyses of UNC93B1 and TLR7 (left)/TLR8 (right) interaction in HEK293T cells. (C) TLR8-immunoprecipitation and western blot analyses of UNC93B1 and TLR8 interaction in HEK293T cells. (D) BioID and western blot analyses of UNC93B1 and TLR7 interaction in RAW 264.7 cells. (E) Immunoblots of TLR7 in endosomes and late endosomes/lysosomes from RAW 264.7 cells expressing the WT or R95L mutant UNC93B1. (F, G) Representative immunofluorescent images (F) and quantification of the colocalization between Tlr7 and Lamp1 or Eea1 in BMDMs from the indicated mice (G) (n = 14-18 cells per group). Data represent mean ± SEM; two-tailed unpaired Student’s *t* test. (H) RNA pull-down and western blot analyses of ssRNA binding affinity of Tlr7 in BMDCs. (I) Graphic model of how R95L mutation affect UNC93B1/TLR axis and downstream signaling.

The weakened interaction did not alter TLR7 trafficking into endosomes, as receptor cleavage, which occurs within endosomes, was unaffected (**Figure 5D**). We also isolated endosomes and lysosomes (late endosomes) from RAW 264.7 cells. Western blot analysis showed comparable levels of TLR7 in both wild-type and UNC93B1 R95L mutant cells (**Figure 5E**). Immunofluorescence microscopy revealed normal colocalization of TLR7 with Eea1 and Lamp1 in mouse BMDMs (**Figure 5, F and G**). These findings indicate that the UNC93B1 R95L mutation does not impact the trafficking of TLR7 from the ER to endosomes or from endosomes to lysosomes, suggesting a trafficking-independent role for UNC93B1.

The weakened interaction between UNC93B1 and TLR7 might alter the receptor conformation and increase the affinity for ligand binding, thereby activating the inflammatory pathway. To explore this, we conducted RNA pull-down experiments using mouse BMDCs. The results revealed that the UNC93B1 R95L significantly enhanced the binding of TLR7 to ssRNA (**Figure 5H**).

## Discussion

Our study characterized a novel homozygous gain-of-function mutation p.R95L in *UNC93B1* leading to early-onset SLE. We also delineate distinct altered inflammatory responses amidst organs and immune cell populations during the SLE development caused by the *UNC93B1* R95L mutation. Limited by the heterogeneity of SLE, preventing SLE patients from sequalae during SLE progression is challenging. Our work enriched the spectrum of SLE pathogenic mutations, which could ameliorate the prognosis and disease development of SLE-like patients through early diagnosis and intervention.

UNC93B1 deficiency was firstly reported in 2006 with immunodeficiency and infections(16). Recently, a series of *UNC93B1* gain-of-function mutations were reported as pathogenic, predisposing to early-onset SLE and arthritis(23–27). These *UNC93B1* gain-of-function mutations exhibit dominant and recessive inheritance, however, some mutations are not rare variants in public database like gnomAD, which is unusual for monogenic disorders caused by gain-of-function mutations. Besides, some mutations, such as T93I, R336C, and V117L, show dosage effects in phenotype presentation in both patients and animal models to different extents. In our study, the *UNC93B1* R95L mutation also displays additive effects on autoimmune pathology and inflammatory response in different tissues. Though parents of patients with UNC93B1 gain-of-function mutations (R95L, E92G, I317M) have not presented with SLE manifestation, current results suggest that gain-of-function *UNC93B1* mutations, no matter mono-allelic or bi-allelic, have potential for SLE development. The variable expressivity and penetrance in the SLE caused by *UNC93B1* gain-of-function mutations implicate a veiled function of UNC93B1 in the innate immune, encouraging further investigation into the mechanism of UNC93B1 in SLE pathogenesis beyond the regulation of TLR signaling.

As unrestricted intracellular nucleic-acid sensing leads to autoimmune or inflammatory diseases(28–31), activation of intracellular TLRs is restrained within cells. TLR7 must be transported to the endosome, where it is cleaved by proteases in the Z-loop region for dimerization. TLR7 has two substrate-binding sites that recognize different substrates. The sequential binding of these substrates induces conformational changes in TLR7, leading to the close dimerization of the C-terminal TIR domain and initiating signal cascades. Subsequently, activated TLR7 is sorted into multivesicular bodies (MVBs) and late lysosomes for degradation, terminating the signal.

UNC93B1 is involved not only in the transport of TLRs to endosomes but also in orchestrating their functions. The D34A mutation in N-terminal domain of UNC93B1 can enhance the response of TLR7 while inhibiting the response of TLR9, leading to systemic lethal inflammation in mice(32). The C-terminal end of UNC93B1 interacts with syntenin-1 to transport TLR7 to the phagosome, thereby terminating TLR7 activation. A mutation in the C-terminal region (PKP530-532) weakens this interaction with syntenin-1, promoting TLR7 activation and resulting in lupus-like symptoms in mice(33). Additionally, studies have shown that the interaction between UNC93B1 and TLR9 in endosomes affects TLR9’s ligand binding and activation. The S282A point mutation disrupts the dissociation of UNC93B1 from TLR9 in endosomes, thereby impairing TLR9 signaling pathway activation in mouse macrophages and dendritic cells(34).

In our study, both humans and mice carrying the *UNC93B1* R95L homozygous mutation exhibited autoimmune and systemic inflammatory phenotypes. Based on our mechanistic research and existing structural studies, we propose the following model (**Figure 5I**): in the absence of ligand binding, TLR7 or TLR8 exists as a pre-dimer with the C-terminal TIR domains far apart and the N-terminal domains unfavorable for ligand binding. Upon ligand binding at the first binding site, the LRR domain undergoes a conformational change that promotes binding at the second site, further driving conformational changes that lead to the aggregation of the C-terminal TIR domains and the initiation of downstream signaling pathways. The two-turn helix H1 (residues 91-97) formed between the TM1 and TM2 transmembrane domains of UNC93B1 directly contacts two loop regions of the TLR7 C-terminal LRR-CT domain, thus restricting TLR7 conformational changes and setting the activation threshold of potentially self-reactive TLR7. The *UNC93B1* R95L mutation weakens the interaction between UNC93B1 and TLR7, enhancing the binding of the LRR domain to the ligand and activating downstream signaling pathways. Recent studies reported that the E92G and T93I mutations also weaken the interaction between UNC93B1 and TLR7, leading to TLR7 hyperactivation, likely through a similar mechanism.

Gain-of-function UNC93B1 mutations affecting innate immune through TLR7/8 but not other endogenous TLRs have been well established in recent research. Our results support this selective activation of TLRs in mutant UNC93B1-mediated inflammation. The interface between UNC93B1 and TLR3 or TLR7 shows no differences, while the crystal structures of TLR7/8 display their distinct oligomerization states in ligand recognition(17, 35), implicating a different mechanism in TLR signaling activation. TLR7 and TLR8 exist as pre-dimers even in the absence of ligand binding, with the regulation of antagonizing binding of UNC93B1 (in a 2:2 pattern) changing the conformation from inactive to active forms. In contrast, TLR3 and TLR9 are monomers that interact with UNC93B1 in a 1:1 pattern without ligand binding. Upon stimulation, UNC93B1 dissociates, allowing the formation of active dimers. This may partially explain why the UNC93B1 R95L mutation specifically promotes the activation of TLR7 and TLR8, but has no effect on the activation of TLR3 and TLR9. These findings indicate the diverse function of UNC93B1 in intracellular nucleic acid homeostasis, suggesting the importance of future research on the dynamic balance of endosomal TLRs during SLE development resulting from activated UNC93B1.

In our results, the *UNC93B1* R95L mutation disrupts the interaction between UNC93B1 and TLR7 as the previously characterized UNC93B1 H412R mutation does, however, UNC93B1 R95L still promotes TLR7/8 signaling. It was reported that the exogenous antigen processing is abolished in *UNC93B1^H412R^* mice(13). Based on the expression pattern of UNC93B1 in protein atlas database, UNC93B1 is absent in T cells. Nevertheless, in our results, T cells from the patient and animal model with the UNC93B1 R95L mutation display activated state in the development of SLE through activation from DCs. Besides, we also observed specific enrichment of immunoglobulin production and SLE signaling in B cells from *Unc93b1^R95L/R95L^* mice. Our results emphasize the importance of antigen presentation and humoral immunity in the immune dysregulation mediated by *UNC93B1* gain-of-function mutations.

To treat SLE caused by TLR7/8 hyperactivation due to UNC93B1 mutations, several approaches may be considered. JAK inhibitors can block the JAK-STAT signaling pathway, reducing inflammation and immune responses driven by cytokines such as IFN-α, which are upregulated by TLR7/8 activation. Hydroxychloroquine (HCQ) inhibits endosomal acidification, thus blocking TLR7/8 activation, and is already widely used in SLE treatment with a good safety profile. Additionally, inhibitors targeting TLR7/8, MyD88, and IRAK1/4 can block the TLR7 signaling pathway with better precision, reducing inflammation with fewer potential off-target effects. These inhibitors, though promising, are still largely undergoing clinical trials and require further evaluation for long-term safety and efficacy.

## Methods

### Sex as a biological variable

Our study examined male and female mice, and sex-dimorphic effects are reported. The sex of human involved in this study was not considered as a biological variable.

### Mice

*Unc93b1* R95L point mutation mice were generated in a C57BL/6J background using CRISPR-Cas9-mediated gene editing technology. Genomic sequence (ENSMUST00000162708.7) was obtained from Ensembl (https://ensembl.org/) and compared to human genes to ascertain sequence conservation. The sgRNA used was 5’-GCCATACTTCACCTCTCTGT-AGG-3’. The donor single-stranded oligonucleotide was: 5’-CAGCCTTGCCGTGAGCTTGTTTAGTTGTTCTGAGCCAGACTGATTAGAGCTCTCTACGATGCTCCCTGTCCCCAGGCCTCCTGCAGATGCAACTGATCCTGCACTATGATGAGACCT AC**CTC**GAGGTGAAGTATGGCAACATGGGGCTGCCGGACATCGATAGCAAGATGCTGAT GGGTATCAACGTGACGCCTATCGCTGCCCTGCTCTACACACCTGTGCTCATCAGGTGCCAAACTTC-3’. The primers designed to amplify the region including the R95L mutation are as follows: Forward, 5’-CTGGGAACGGGAGTCTTGT-3’; Reverse, 5’-AACAAGCAAGGCCTCTCTGC-3’.

Both male and female mice were used and their genders are indicated in the figures or figure legends. Within genotypes, mice were randomly allocated in all experiments. Data collection and analysis were not performed blind to the conditions of the experiments, except for histopathology. No animals or data points were excluded except contaminated samples.

### Whole exome sequencing and Sanger sequencing

Genomic DNA from the patient and family members were isolated from peripheral blood using Maxwell RSC Whole Blood DNA Kit (Promega, AS1520). Whole exome sequencing (WES) and data analysis were performed as previously described(36, 37). ANNOVAR (2019Oct24) was used to annotate variants. Variants that appeared in gnomAD, Kaviar, dbSNP, and an in-house database were filtered out. Variants were further filtered by homozygous inheritance. Sanger sequencing was used to confirm the *UNC93B1* variant identified by WES.

### Cell preparation, culture, and stimulation

RAW 264.7, THP-1 and HEK293T cell lines were obtained from the American Type Culture Collection. All cell lines tested negative for Mycoplasma contamination. PBMCs were separated by Lymphocyte Separation Medium (MP, 50494) according to the manufacturer’s instructions. BMDMs were differentiated for 6-8 days in complete medium containing 50 ng/mL M-CSF (PeproTech, 315-02). BMDCs were differentiated for 7 days in complete medium containing 20 ng/mL GM-CSF (PeproTech, 315-03). For UNC93B1 overexpressing stable cell lines, a coding sequence expressing UNC93B1 and BirA* biotin ligase fusion protein (UNC93B1-BirA*) was cloned into the retroviral MSCV-puro vector. THP-1 and RAW 264.7 cells were transduced with retroviruses expressing UNC93B1-BirA* and subjected to puromycin selection.

RAW 264.7, HEK293T and BMDMs were grown in Dulbecco’s Modified Eagle Medium (DMEM, Gibco) supplemented with 10% FBS (Noverse) and penicillin/streptomycin (Gibco). THP-1, PBMCs and BMDCs were grown in RPMI-1640 (Gibco) supplemented with 10% FBS and penicillin/streptomycin.

R848 (Resiquimod, Sigmaaldrich, SML0196), a dual TLR7 and TLR8 synthetic agonist was used to stimulate HEK293T (5 μg/mL), RAW 264.7 (25 ng/mL, 100 ng/mL), BMDCs (25 ng/mL), and THP-1 (100 ng/mL) for the indicated amount of time. ssRNA40 (InvivoGen, tlrl-lrna40), the natural ligand of the TLR7 and TLR8 was used to stimulate BMDCs (0.1 μM, 1 μM), BMDMs (1 μM) for the indicated amount of time. CL307 (InvivoGen, tlrl-c307) a specific TLR7 agonist was used to stimulate RAW 264.7 (50 ng/mL) for the indicated amount of time. Guanosine (Sigmaaldrich, G6264) was used to stimulate RAW 264.7 (200 μM) for the indicated amount of time. TL8-506, a specific agonist for TLR8, was used to stimulate THP-1 (0.1 μg/mL, 1 μg/mL) for the indicated amount of time. Polyinosinic-polycytidylic acid (Poly(I:C), Sigmaaldrich, P9582) a synthetic analog of double-stranded RNA (dsRNA) was used to stimulate HEK293T (50 μg/mL), RAW 264.7 (10 μg/mL), BMDCs (10 μg/mL), and THP-1 (10 μg/mL) for the indicated amount of time. LPS (Sigmaaldrich, L6529) was used to stimulate RAW 264.7 (0.1 μg/mL) and THP-1 (0.1 μg/mL) for the indicated amount of time. Class A CpG oligonucleotide ODN2216 (InvivoGen, tlrl-2216) was used to stimulate HEK293T (5 μM) for the indicated amount of time. Class B CpG oligonucleotide ODN2006 (InvivoGen, tlrl-2006) was used to stimulate THP-1 (5 μM) for the indicated amount of time. Class A CpG oligonucleotide ODN1585 (InvivoGen, tlrl-1585) was used to stimulate BMDCs (0.5 μM, 5 μM) and RAW 264.7 (0.5 μM, 5 μM) for the indicated amount of time. Class B CpG oligonucleotide ODN1668 (InvivoGen, tlrl-1668) was used to stimulate BMDMs (0.5 μM, 5 μM), BMDCs (0.5 μM, 5 μM) and RAW 264.7 (0.5 μM, 5 μM) for the indicated amount of time. Chloroquine (CQ, Selleck, S6999), an autophagy/lysosome inhibitor, was used to treat RAW 264.7 (50 μM) for the indicated amount of time. Enpatoran (M5049, Selleck, S9931) hydrochloride a dual TLR7/8 inhibitor, was used to treat RAW 264.7 (10 μM) for the indicated amount of time.

### Expression plasmids and antibodies

Human TLR3, TLR7, TLR8, TLR9 and wild-type UNC93B1 plasmids were constructed by PCR amplification of UNC93B1 from the cDNAs of the THP-1 cell line, and then cloned into the pLenti vector or MSCV-puro vectors made in-house, and the mutant plasmids were constructed by site-directed mutagenesis.

For western blotting experiments, TLR7 (5632), TLR8 (11886), TLR9 (13674), and GAPDH (5174) antibodies were purchased from Cell Signaling Technology. Flag (F3165) antibody was from Sigma aldrich. UNC93B1 (PA5-20510) antibody was from Invitrogen. LAMP1 (ab25630) antibody was from Abcam.

For immunofluorescence experiments, TLR7 (5632) and EEA1(3288) antibodies were purchased from Cell Signaling Technology. LAMP1 (ab25630) antibody was purchased from Abcam.

For flow cytometry experiments, V500 Rat Anti-Mouse CD45(30-F11) (561487), BV421 Hamster Anti-Mouse CD3e(145-2C11) (562600), FITC Rat Anti-Mouse CD4(RM4-5) (553046), APC-Cy7 Rat Anti-Mouse CD8a(53-6.7) (557654), APC Rat Anti-Mouse CD19(1D3) (550992), PE Rat Anti-Mouse CD138(281-2) (553714), BV605 CD317 (BST2) (747606), PE-Cy7 Rat Anti-Mouse CD45R/B220(RA3-6B2) (552772), BUV496 CD11b (749864), BV750 F4/80 (747295), RB780 Ly-6C (755871), BUV395 I-A, I-E (569244), BUV805 CD44 (741921), BUV563 CD62L (741230), BV786 Rat Anti-Mouse CD25(PC61) (564023), BV650 Rat Anti-Mouse IL-17A (TC11-18H10) (564170), BUV737 Rat Anti-Mouse CD21/CD35(7G6) (612810), RB545 CD23 (756344), BV480 CD95 (746755), BUV615 CD49b (751052), BV711 CD279 (PD-1) (744547), PE-CF594 Rat Anti-Mouse CD185 (CXCR5)(2G8) (562856), and R718 Ly-6G (567039) antibodies were purchased from BD Biosciences. PE-CYN5 FOXP3 (15-5773-82), PE-CYN5 FOXP3 (15-5773-82), and PERCPEF710 BCL-6 (46-5453-82) antibodies were purchased from Invitrogen. Brilliant Violet 570 anti-mouse CD11c (117331) and APC anti-mouse TNF (506308) antibodies were purchased from Biolegend.

### Dual-luciferase reporter assay

The NF-κB firefly reporter plasmid (pGL4.32 [luc2P/NF-κB-RE/Hygro] and the Renilla luciferase expression plasmid (pRLCMV-Renilla) were co-transfected along with TLRs and UNC93B1 expression plasmids into HEK293 cells. After 24 hours of transfection and indicated treatment, cells were harvested, and luciferase activity was measured using the Dual Luciferase Reporter Assay Kit (Vazyme, DL101-01). Data were calculated as fold induction by normalizing firefly luciferase activity to Renilla luciferase activity.

### RNA extraction and quantitative PCR

TRIzol reagent (Invitrogen, 15596026) and RNeasy Mini kit (Qiagen, 74104) were applied to extract total RNA from cultured cells and murine tissues. One microgram of RNA was reverse-transcribed using the PrimeScript RT Reagent Kit with gDNA Eraser (Perfect Real Time) (Takara, RR047A). Gene expression analyses were performed using 2X Universal SYBR Green Fast qPCR Mix (ABclonal, RK21203) on ROCHE 480II. Relative mRNA expression was analyzed by the ΔΔ*C*_t_ method and normalized to *ACTB* (for human samples) or *Actb* (for mouse samples).

### RNA sequencing

RNA sequencing libraries were generated using the NEBNext Ultra RNA Library Prep Kit for Illumina (New England Biolabs) following the manufacturer’s protocol. The libraries were then sequenced on an Illumina NovaSeq platform to obtain high-throughput sequencing data. The resulting reads were mapped to the reference genome, and featureCounts was utilized to quantify the number of reads mapped to each gene. For differential expression analysis, the DESeq2 (version 1.44.0) and clusterProfiler (version 4.12.0) R package was employed, which involved normalization of read counts, statistical testing, and identification of differentially expressed genes between the experimental conditions. Gene set scores in differential expression analysis of RNA sequencing data are summing of z-scores of each gene in the gene set. Z-score of each gene in the gene set is calculated relative to the mean and standard deviation of Healthy controls. Gene sets from msigDB (https://www.gsea-msigdb.org/gsea/msigdb) are used for analysis.

### Single-cell RNA sequencing

For human PBMCs, 8,000-10,000 single cells for each sample were captured and barcoded with a 10x Genomics Chromium console, while the library of about 10,000 cells from murine spleen samples were constructed by MGI DNBelab C-TaiM4. The barcoded complementary DNA was amplified and sequenced by Illumina Novaseq and MGI DNBSEQ-T7 for human PBMC and murine samples, respectively. Novaseq sequencing data was processed with cellranger (10X Genomics, version 6.1.1), while DNGSEQ-T7 sequencing data was processed with DNBC4tools (MGI, version 2.1.1). Downstream quality control, processing and differential expression analysis were performed by Seurat package (version 4.2.0) and harmony package (version 1.2.0) in R.

### Flow cytometry analysis

The mice spleen single-cell suspensions were prepared using the plunger end of the syringe and filtered using 100 µm and 40 µm strainers with 2% FBS in PBS. The collected filtrates were centrifuged at 350 g for 5 min. The cell pellet was suspended in 5 mL RBC lysis buffer, incubated for 4 min at 25 °C and then mixed with 10 mL of PBS with 2% FBS. The splenocytes were resuspend in PBS after centrifugation at 350 g for 5 min. Whole blood cells were collected from heart blood and incubated with 10 mL RBC lysis buffer for 5 min at 25 °C, and then mixed with 20 mL PBS with 2% FBS. The whole blood cells were resuspended in PBS after centrifugation at 350 g for 5 min.

For flow cytometry analysis, 1*10^6^ cells were incubated with Fixable Viability Stain 440UV (BD Biosciences, 566332) for 15 min at room temperature protected from light. Washed twice with fluorescence-activated cell sorting (FACS) buffer (0.5% BSA in PBS) and stained with cell-surface markers for 1 hours on ice. For fixation and permeabilization, 1.6% PFA and 40% methanol were used. The cells were then washed twice and stained with intracellular antibodies for 1 hours on ice. All events were acquired on Cytek Aurora and analyzed by SpectroFlo and FlowJo_v.10.8.1. Gating strategy is shown in Supplemental Figure 6.

### ELISA and CBA

Levels of cytokines IL-6, IFN-α and chemokine IP-10 in human serum were determined by Cytometric Bead Array (BD Bioscience). All data were analyzed by FCAPArray V3 software (BD Biosciences). Macrophage colony-stimulating factor (M-CSF) (MULTISCIENCES, EK1144), Granulocyte colony-stimulating factor (G-CSF) (MULTISCIENCES, EK169) concentrations in human serum were measured by enzyme-linked immunosorbent assay (ELISA) kit.

The ELISA kits used for Cxcl1 (EK286), Il-6 (EK206), CRP (EK294) quantification in murine plasma were purchased from Multisciences. ANA (ml002245), Il-1β (ml098416) and C3 (ml002033) quantification in murine plasma were purchased from Mlbio.

For detecting autoantibodies in murine plasma, coating antigens (1 μg DNA or RNA) were diluted in Coating Buffer and added to an ELISA plate, then incubated overnight at 4 °C. The plate was blocked with Diluent Buffer for 1 hour at room temperature. Samples and standards were diluted in Diluent Buffer and incubated for 2 hours at room temperature. Detection antibodies were added and incubated for 1 hour, followed by HRP conjugate (Beyotime, A0216) for 30 minutes, both at room temperature. TMB substrate (Beyotime, P0209) was added and incubated for 30 minutes, then stop solution (Beyotime, P0215) was added. Absorbance was measured at 450 nm using a microplate reader.

### Western blotting and Immunoprecipitation

Cells were lysed in ice-cold lysis buffer ( 50 mM Tris–HCl, pH 8.0, 150 mM NaCl, 5 mM EDTA, 1% digitonin) supplemented with Complete Mini EDTA-free (4693159001, Roche) and PhosSTOP (4906837001, Roche). Protein concentration was measured by a BCA protein assay kit (23225, Thermo Fisher). Immunoblotting was conducted as described previously with specific antibodies(38, 39).

For immunoprecipitation, cell lysates were combined with immunomagnetic beads for 4 hours. Then, immunocomplexes were washed five times using the lysis buffer and subjected to western blot analysis.

The proximity-labeling BioID approach, as previously described(40), was used to detect the interactions between UNC93B1 and TLR7. RAW 264.7 cells stably expressing UNC93B1 (WT)-BirA* /UNC93B1 (R95L)-BirA* were incubated for 24 hours in complete media supplemented with or without 20 μM biotin. After three washes with PBS, cells were lysed in lysis buffer (50 mM Tris-Cl, 137 mM NaCl, 1 mM EDTA, 1% Triton X-100, 10% Glycerol) supplemented with Complete Mini EDTA-free (4693159001, Roche). Lysed cells were centrifuged under 14,000 g for 10 min at 4 °C. Supernatants were then incubated with 20 μL streptavidin agarose beads for 24 h at 4 °C. Beads were pelleted by centrifugation at 2,000 rpm for 2 min and eluted with Elute Buffer (1.5x SDS protein sample buffer containing 1 mM biotin) for western blot analysis.

### Immunofluorescence

For immunofluorescence (IF) staining of murine kidney tissue cryosections, the sections were first fixed and permeabilized, followed by blocking with 10% normal goat serum (NGS, Beyotime, C0265) in PBS for 1 hour. Mouse antibodies diluted in 10% NGS/PBS (IgG 1:200; C3 1:200) were applied to the sections and incubated overnight at 4°C. Secondary antibodies diluted at 1:500 in 10% NGS/PBS were then incubated with the sections at room temperature for 1 hour.

For IF staining of cultured cells, cells were plated onto glass coverslips and allowed to settle overnight. Cells were fixed with methanol for 5 min at –20 °C and permeabilized with 100 µM digitonin-PBS for 5 min at RT. After washing 3 times with PBS, cells were blocked in 10% NGS (Beyotime, C0265) in PBS for 30min. Slides were stained in blocking buffer with primary antibodies overnight at 4 °C. After washing 3 times with PBS, slides were incubated for 1 hour with secondary antibodies in blocking buffer at RT. After washing 3 times with PBS, slides were mounted using Mounting Medium with DAPI (ab104139, abcam). Images were acquired and quantified by ZEISS LSM 900.

### Immunohistochemistry

Freshly dissected murine spleens, kidneys, livers and lungs were fixed in 4% paraformaldehyde (Sigma-Aldrich, P6148) for 16 hours at 4 °C, embedded with paraffin and stained with hematoxylin/eosin (H&E) or periodic acid-Schiff (PAS). The tissue slides were scanned using Aperio ScanScope XT scanner (Leica). Glomerular size was analyzed in a blinded manner using NDP.view 2. Glomerular size was determined from an area with at least 10 glomeruli per kidney section from at least 3 mice in each group.

### Author contributions

QZ, PT and XY designed the study, directed, and supervised the research. XH, RW, SO, and QW contributed equally. PT, RW, QW, WD, YZ, OX and HY performed experiments. XH analyzed the data. QZ and SO enrolled the patient, collected, and interpreted clinical information. QZ, PT, XH, and RW wrote the manuscript, with input from others. All authors contributed to the review and approval of the manuscript.

## Data Availability

All data produced in the present study are available upon reasonable request to the authors

## Acknowledgments

We thank the patient and the unaffected controls for their support during this research study. We thank the core facility of the Life Sciences Institute, Zhejiang University and core facilities of Zhejiang University Medical Center/Liangzhu laboratory for technical assistance. The works of QZ were supported by grants 82225022, 32141004 and 32321002 from The National Natural Science Foundation of China. The works of X.Y. were supported by the Major Program of the National Natural Science Foundation of China (82394424), the National Natural Science Foundation of China (82471844), the Hundred-Talent Program of Zhejiang University and Leading Innovative and Entrepreneur Team Introduction Program of Zhejiang (2021R01012). The works of PT were supported by grants 82201926 and 82471814 from The National Natural Science Foundation of China, BX2021259 and 2021M702852 from China Postdoctoral Science Foundation. The works of RW were supported by grant 82300893 from The National Natural Science Foundation of China.

## Statistics

Statistical analyses were performed using GraphPad Prism 8. All values were as mean ± SEM and calculated from the average of at least three independent biological replicates unless specifically stated. Statistical differences were evaluated using the Student’s *t* test (unpaired and two-tailed) for comparisons between two groups or ANOVA for comparisons of more than two groups. In all tests, a 95% confidence interval was used, for which *P* < 0.05 was considered a significant difference. *P* values obtained in next-generation sequencing were adjusted by FDR using R Software (R version 4.4.1).

## Study approval

This study involves human participants and was approved by the institutional review boards of the Children’s Hospital of Zhejiang University School of Medicine in China. Participants provided written informed consent to participate in the study before taking part.

Mice were bred and maintained in specific-pathogen-free conditions at the Laboratory Animal Center of Zhejiang University, with normal diet, 12 h light/dark cycle, 20-26 °C and humidity of 40-70%. Experimentation was approved by the Institutional Animal Care and Use Committee of Zhejiang University.

## Data and materials availability

All data are available in the main text or the Supplemental materials.

## Figures

**Supplemental Figure 1.**
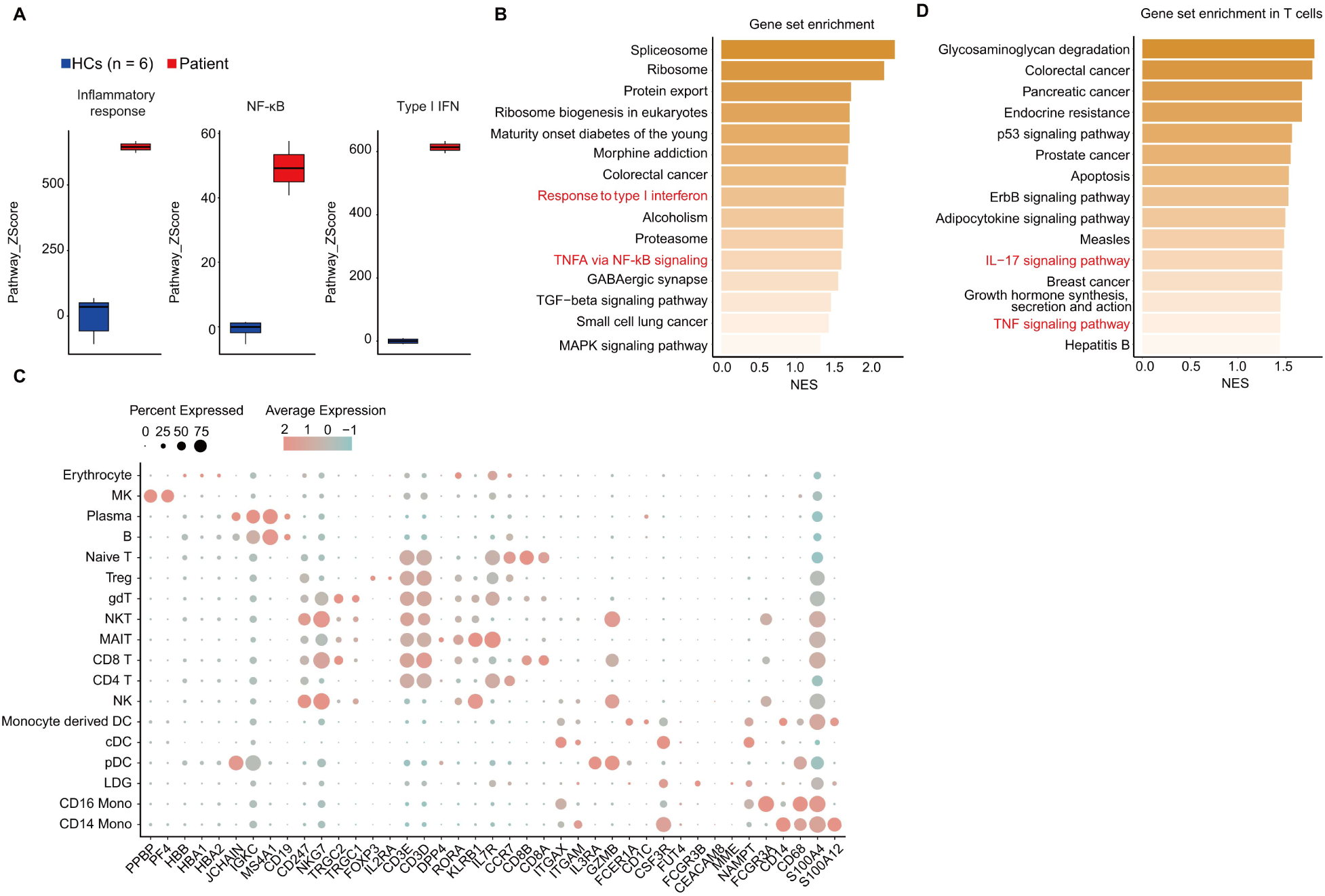
Inflammatory signaling activated in the patient with UNC93B1 R95L mutation. (A) Comparison of gene set score of inflammatory response genes in msigDB and NF-κB and type I IFN response genes between patient and healthy controls (HCs, n = 3). (B) Top 15 enrichment of upregulated pathways of differential expressed gene in patient based on GSEA. (C) Marker genes used for cluster annotation in human PBMCs single-cell RNA sequencing. (D) Top 15 enrichment of upregulated pathways of differential expressed gene in patient’s T cells based on GSEA.

**Supplemental Figure 2.**
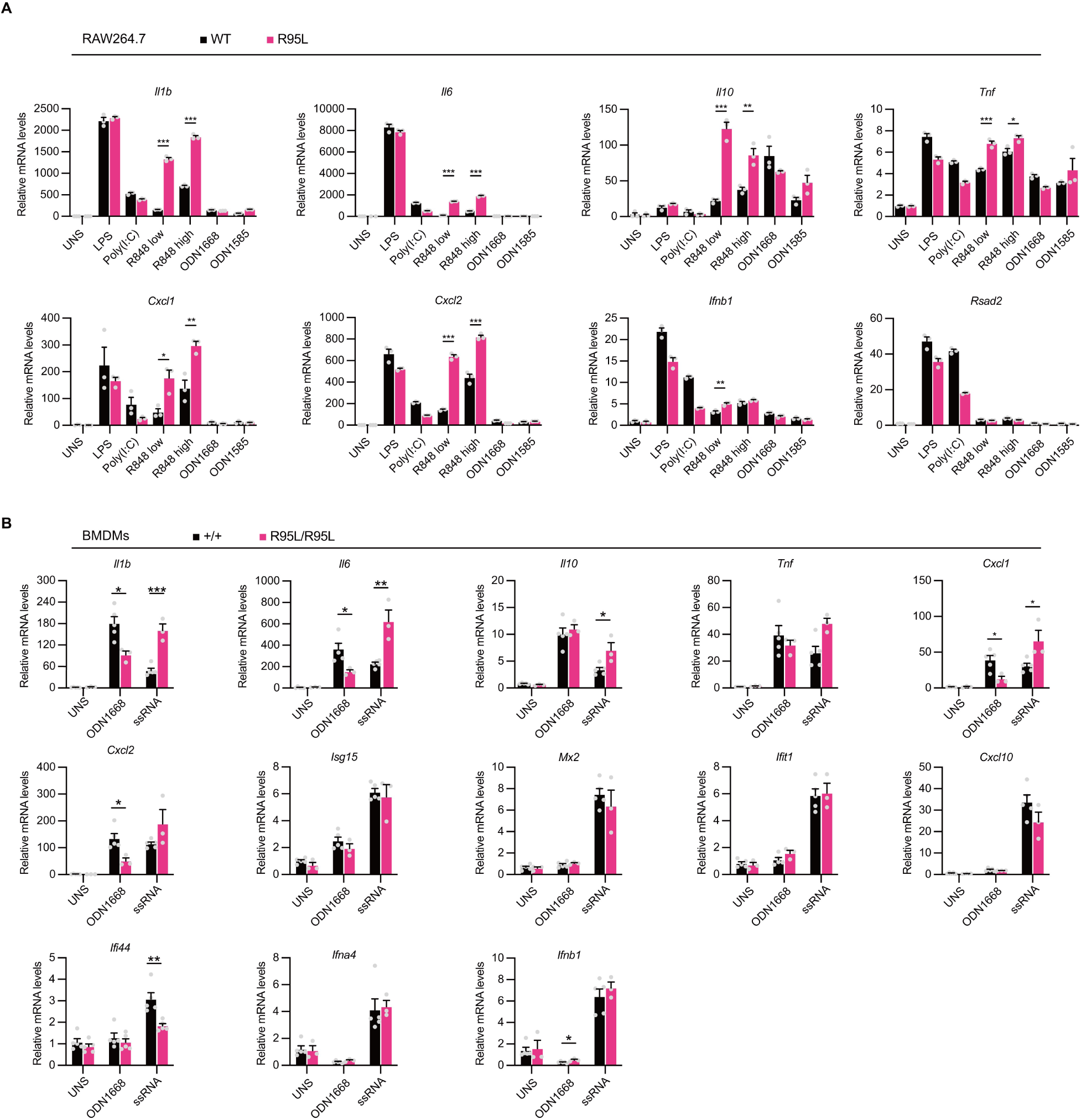
UNC93B1 R95L mutation promotes inflammation through TLR7/TLR8 activation. (A) qPCR analysis of NF-κB and type I IFN pathways related genes transcription in RAW 264.7 cells treated with various TLRs agonists (n = 3 biological replicates). Data represent mean ± SEM; *, *P* < 0.05; **, *P* < 0.01; ***, *P* < 0.001; two-tailed unpaired Student’s *t* test. (B) qPCR analysis of NF-κB and type I IFN pathways related genes transcription in BMDMs from indicated mice treated with various TLRs agonists (n = 3-5 mice per group). Data represent mean ± SEM; *, *P* < 0.05; **, *P* < 0.01; ***, *P* < 0.001; two-tailed unpaired Student’s *t* test.

**Supplemental Figure 3.**
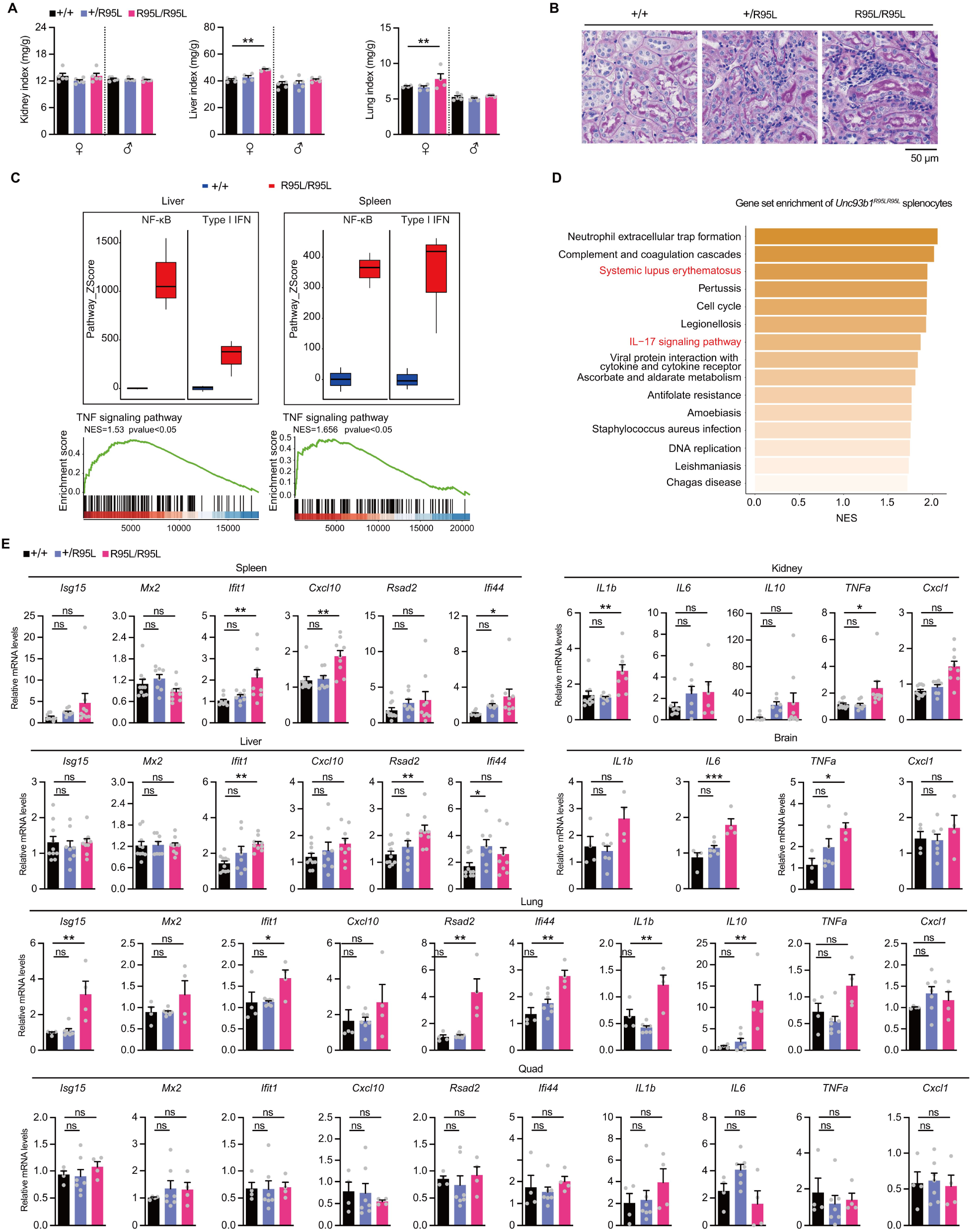
Mice with *Unc93b1^R95L^* mutation develop systemic inflammation and autoimmune pathology. (A) Normalized kidney, liver, lung weight to body weight of 12-wk-old mice (female, n = 4 per group; male, n = 5 per group). Data represent mean ± SEM; **, *P* < 0.01; one-way ANOVA. (B) PAS staining of the kidney from indicated mice. (C) Comparison of gene set score of NF-κB and type I IFN response genes and GSEA in TNF signaling pathway of liver and spleen tissues from the *Unc93b1^+/+^*and *Unc93b1^R95L/R95L^*mice. NES, normalized enrichment score. (D) Top 15 enrichment of upregulated pathways of differential expressed gene between the *Unc93b1^+/+^* and *Unc93b1^R95L/R95L^* mice based on GSEA. NES, normalized enrichment score. (E) qPCR analysis of NF-κB and type I IFN pathways related genes in the spleen, kidney, liver, brain, lung and quadricep from the indicated mice (spleen, kidney and liver, n = 8-10 per group; brain, lung and quadricep, n = 4-7 per group). Data represent mean ± SEM; *, *P* < 0.05; **, *P* < 0.01; ***, *P* < 0.001; one-way ANOVA.

**Supplemental Figure 4.**
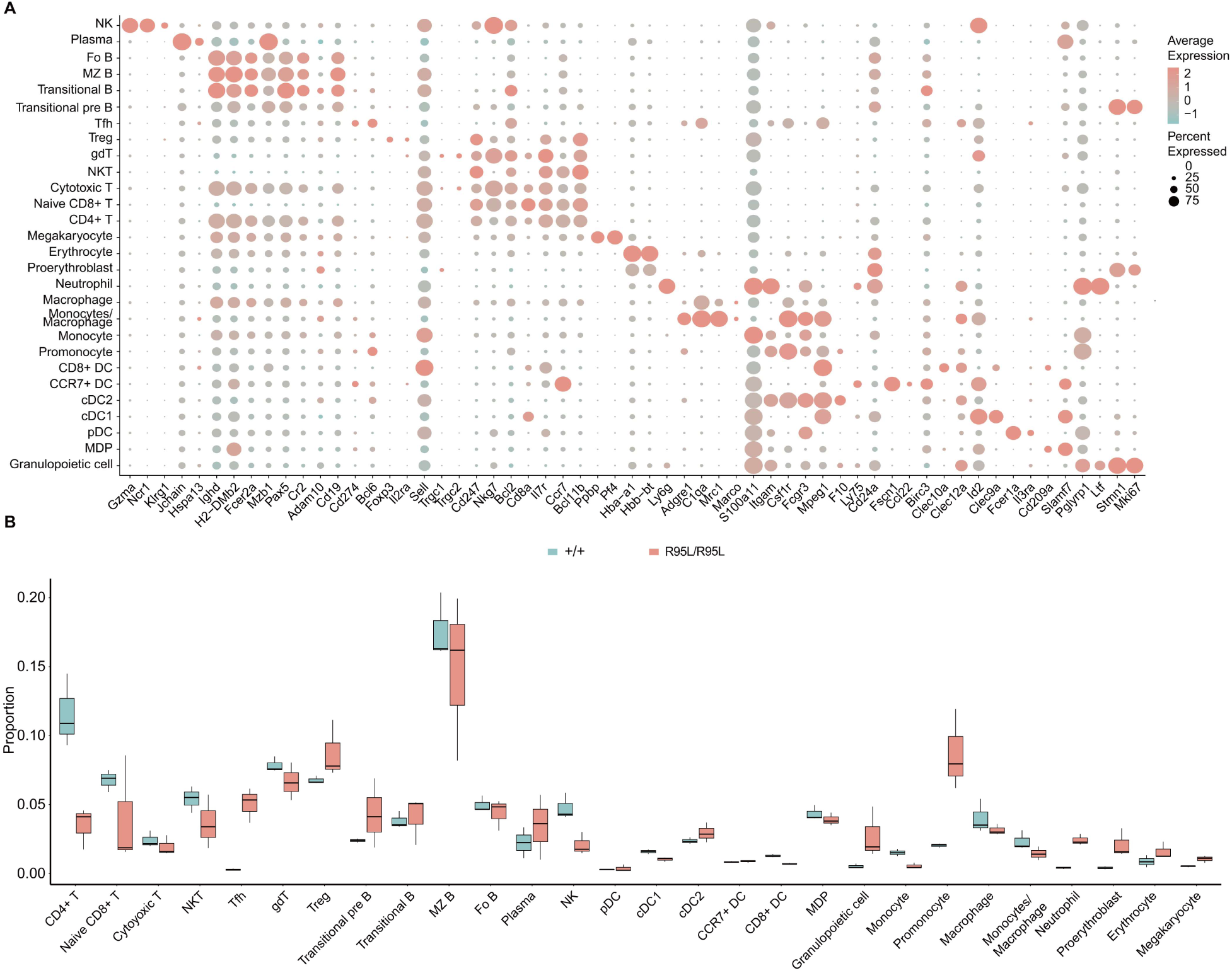
Single-cell sequencing analysis of murine splenocytes. (A) Marker genes used for cluster annotation in murine splenocytes single-cell RNA sequencing. (B) Cell ratio of different cell clusters among murine splenocytes based on single-cell sequencing from the *Unc93b1^+/+^* and *Unc93b1^R95L/R95L^* mice. NKT, natural killer T cell; gdT, gamma-delta T cell; Treg, regulatory T cell; MZ B, marginal zone B cell; Fo B, follicular B cell; DC, dendritic cell; MDP, myeloid dendritic progenitor.

**Supplemental Figure 5.**
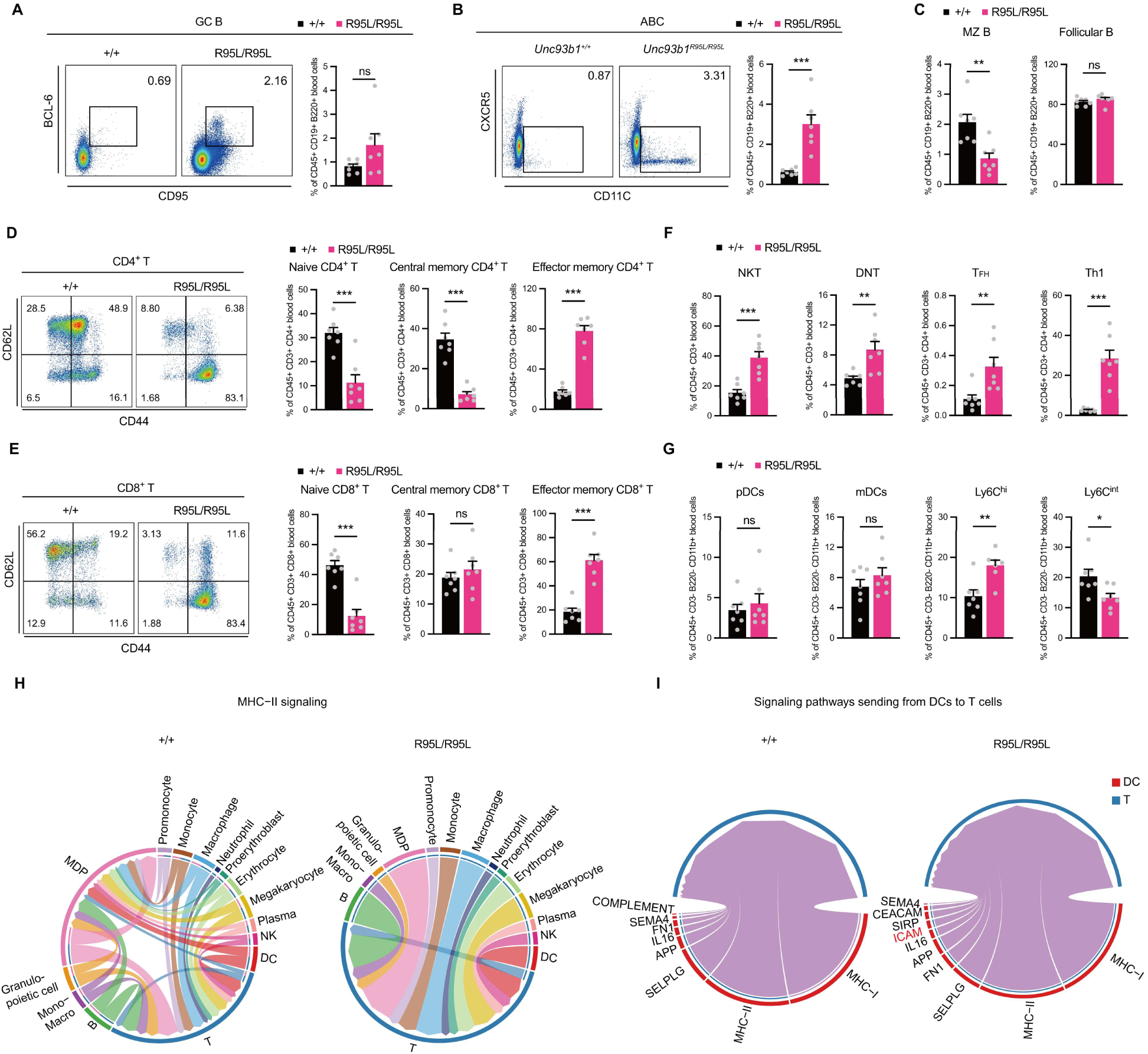
Effects of *Unc93b1^R95L^* mutation on immune cell populations in mice. (A-C) germinal center B cells (GC B) (A), age-associated B cells (ABC) (B), marginal zone B cells (MZ B) and follicular B cells (C) distribution in peripheral blood of *Unc93b1^R95L/R95L^* and *Unc93b1^+/+^*mice are indicated by FACS plots and percentages (n = 7 mice per group). Data represent mean ± SEM; **, *P* < 0.01; ***, *P* < 0.001; two-tailed unpaired Student’s *t* test. (D, E) Peripheral blood CD4^+^ T cells (D) and CD8^+^ T cells (E) activation are indicated by FACS plots and percentages (n = 7 mice per group). Data represent mean ± SEM; ***, *P* < 0.001; two-tailed unpaired Student’s *t* test. (F) Flow cytometric analysis of peripheral blood T cell populations from indicated mice (n = 7 mice per group). Data represent mean ± SEM; **, *P* < 0.01; ***, *P* < 0.001; two-tailed unpaired Student’s *t* test. (G) Flow cytometric analysis of peripheral blood pDCs, mDCs and monocytes populations from indicated mice (n = 7 mice per group). Data represent mean ± SEM; *, *P* < 0.05; **, *P* < 0.01; two-tailed unpaired Student’s *t* test. (H) Cell communication of MHC-II signaling in splenocytes from the *Unc93b1^+/+^* and *Unc93b1^R95L/R95L^*mice. NKT, natural killer T cell; gdT, gamma-delta T cell; Treg, regulatory T cell; MZ B, marginal zone B cell; Fo B, follicular B cell; DC, dendritic cell; MDP, myeloid dendritic progenitor. (I) Signaling received by T cells from DCs in splenocytes from the *Unc93b1^+/+^* and *Unc93b1^R95L/R95L^* mice.

**Supplemental Figure 6.**
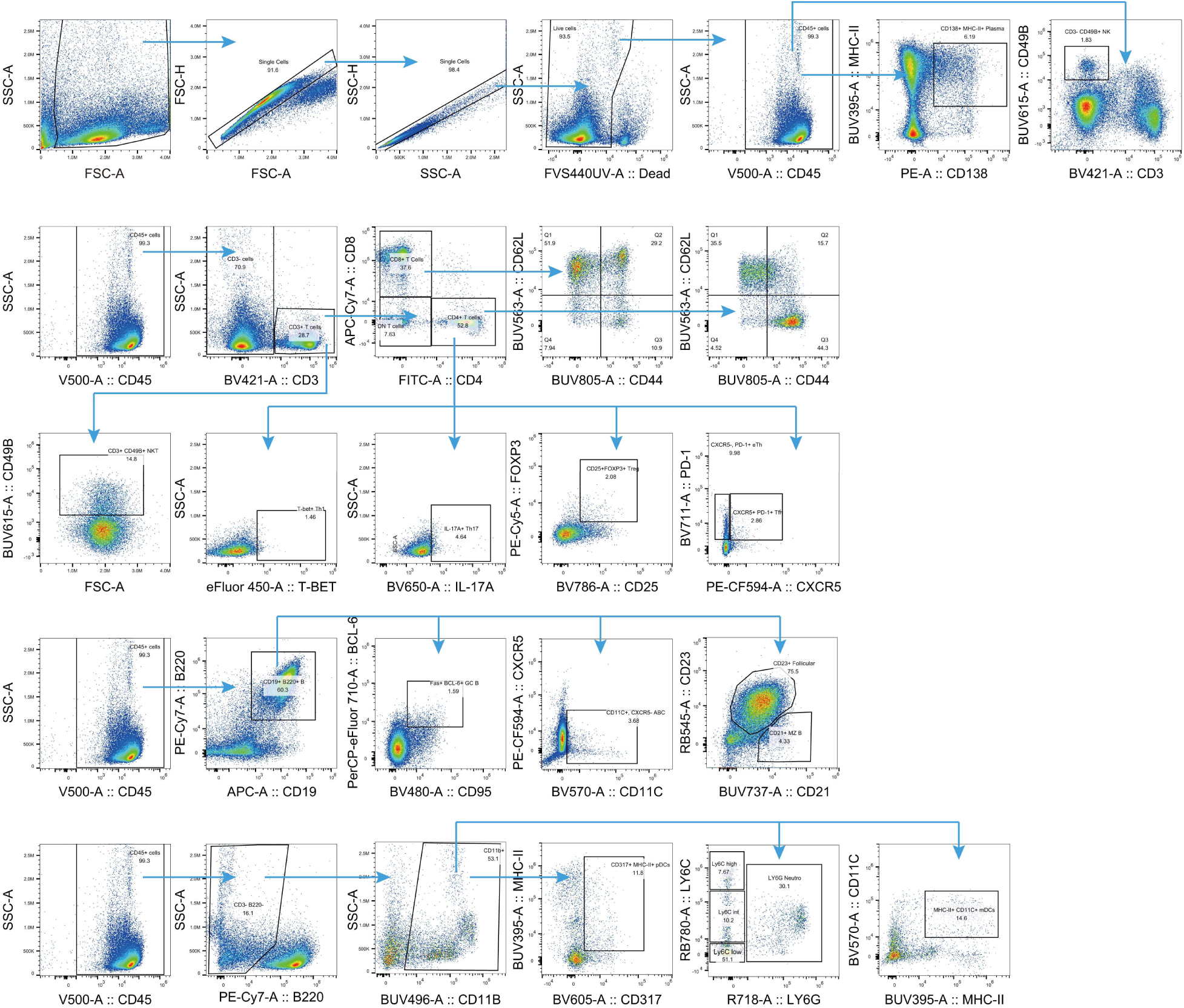
Gating strategies. Representative gating strategies for germinal center B cells (GC B), age-associated B cells (ABC), marginal zone B cells (MZ B), and follicular B cells, plasma cells, CD4+ T cells, CD8+ T cells, NKT, DNT, Treg, Tfh, eTh, Th17, NK, pDCs, mDCs and monocytes in spleen or peripheral blood. These strategies used for data presented in Figure 4 and Supplemental Figure 5.

## References

1. Omarjee O, et al. Monogenic lupus: Dissecting heterogeneity. Autoimmun Rev. 2019;18(10):102361.

2. Demirkaya E, et al. New Horizons in the Genetic Etiology of Systemic Lupus Erythematosus and Lupus-Like Disease: Monogenic Lupus and Beyond. J Clin Med. 2020;9(3).

3. O’Neill LA, and Bowie AG. The family of five: TIR-domain-containing adaptors in Toll-like receptor signalling. Nat Rev Immunol. 2007;7(5):353–64.

4. Fitzgerald KA, and Kagan JC. Toll-like Receptors and the Control of Immunity. Cell. 2020;180(6):1044–66.

5. Gantier MP, et al. TLR7 is involved in sequence-specific sensing of single-stranded RNAs in human macrophages. J Immunol. 2008;180(4):2117–24.

6. Hornung V, et al. Quantitative expression of toll-like receptor 1-10 mRNA in cellular subsets of human peripheral blood mononuclear cells and sensitivity to CpG oligodeoxynucleotides. J Immunol. 2002;168(9):4531–7.

7. van der Made CI, et al. Presence of Genetic Variants Among Young Men With Severe COVID-19. Jama. 2020;324(7):663–73.

8. Brown GJ, et al. TLR7 gain-of-function genetic variation causes human lupus. Nature. 2022;605(7909):349-56.

9. David C, et al. Interface Gain-of-Function Mutations in TLR7 Cause Systemic and Neuro-inflammatory Disease. J Clin Immunol. 2024;44(2):60.

10. Stremenova Spegarova J, et al. A de novo TLR7 gain-of-function mutation causing severe monogenic lupus in an infant. J Clin Invest. 2024;134(13).

11. Cervantes JL, et al. TLR8: the forgotten relative revindicated. Cell Mol Immunol. 2012;9(6):434–8.

12. Aluri J, et al. Immunodeficiency and bone marrow failure with mosaic and germline TLR8 gain of function. Blood. 2021;137(18):2450–62.

13. Tabeta K, et al. The Unc93b1 mutation 3d disrupts exogenous antigen presentation and signaling via Toll-like receptors 3, 7 and 9. Nat Immunol. 2006;7(2):156–64.

14. Itoh H, et al. UNC93B1 physically associates with human TLR8 and regulates TLR8-mediated signaling. PLoS One. 2011;6(12):e28500.

15. Kim YM, et al. UNC93B1 delivers nucleotide-sensing toll-like receptors to endolysosomes. Nature. 2008;452(7184):234–8.

16. Casrouge A, et al. Herpes simplex virus encephalitis in human UNC-93B deficiency. Science. 2006;314(5797):308–12.

17. Ishida H, et al. Cryo-EM structures of Toll-like receptors in complex with UNC93B1. Nat Struct Mol Biol. 2021;28(2):173–80.

18. Aringer M, et al. 2019 European League Against Rheumatism/American College of Rheumatology Classification Criteria for Systemic Lupus Erythematosus. Arthritis Rheumatol. 2019;71(9):1400–12.

19. Postal M, et al. Type I interferon in the pathogenesis of systemic lupus erythematosus. Curr Opin Immunol. 2020;67:87–94.

20. Reizis B. Plasmacytoid Dendritic Cells: Development, Regulation, and Function. Immunity. 2019;50(1):37–50.

21. Granot T, et al. Dendritic Cells Display Subset and Tissue-Specific Maturation Dynamics over Human Life. Immunity. 2017;46(3):504–15.

22. Shin KS, et al. Monocyte-Derived Dendritic Cells Dictate the Memory Differentiation of CD8(+) T Cells During Acute Infection. Front Immunol. 2019;10:1887.

23. Rael VE, et al. Large-scale mutational analysis identifies UNC93B1 variants that drive TLR-mediated autoimmunity in mice and humans. J Exp Med. 2024;221(8).

24. David C, et al. Gain-of-function human UNC93B1 variants cause systemic lupus erythematosus and chilblain lupus. J Exp Med. 2024;221(8).

25. Al-Azab M, et al. Genetic variants in UNC93B1 predispose to childhood-onset systemic lupus erythematosus. Nat Immunol. 2024;25(6):969–80.

26. Mishra H, et al. Disrupted degradative sorting of TLR7 is associated with human lupus. Sci Immunol. 2024;9(92):eadi9575.

27. Wolf C, et al. UNC93B1 variants underlie TLR7-dependent autoimmunity. Sci Immunol. 2024;9(92):eadi9769.

28. Rodero MP, et al. Type I interferon-mediated autoinflammation due to DNase II deficiency. Nat Commun. 2017;8(1):2176.

29. Al-Mayouf SM, et al. Loss-of-function variant in DNASE1L3 causes a familial form of systemic lupus erythematosus. Nat Genet. 2011;43(12):1186–8.

30. Crow YJ, et al. Mutations in the gene encoding the 3’-5’ DNA exonuclease TREX1 cause Aicardi-Goutières syndrome at the AGS1 locus. Nat Genet. 2006;38(8):917–20.

31. Peng J, et al. Clinical Implications of a New DDX58 Pathogenic Variant That Causes Lupus Nephritis due to RIG-I Hyperactivation. J Am Soc Nephrol. 2023;34(2):258–72.

32. Fukui R, et al. Unc93B1 restricts systemic lethal inflammation by orchestrating Toll-like receptor 7 and 9 trafficking. Immunity. 2011;35(1):69–81.

33. Majer O, et al. UNC93B1 recruits syntenin-1 to dampen TLR7 signalling and prevent autoimmunity. Nature. 2019;575(7782):366–70.

34. Majer O, et al. Release from UNC93B1 reinforces the compartmentalized activation of select TLRs. Nature. 2019;575(7782):371–4.

35. Tanji H, et al. Structural reorganization of the Toll-like receptor 8 dimer induced by agonistic ligands. Science. 2013;339(6126):1426–9.

36. Zhou Q, et al. Early-onset stroke and vasculopathy associated with mutations in ADA2. N Engl J Med. 2014;370(10):911–20.

37. Zhou Q, et al. Loss-of-function mutations in TNFAIP3 leading to A20 haploinsufficiency cause an early-onset autoinflammatory disease. Nat Genet. 2016;48(1):67–73.

38. Tao P, et al. A dominant autoinflammatory disease caused by non-cleavable variants of RIPK1. Nature. 2020;577(7788):109-14.

39. Wang Y, et al. Identification of an IL-1 receptor mutation driving autoinflammation directs IL-1-targeted drug design. Immunity. 2023;56(7):1485–501.e7.

40. Roux KJ, et al. A promiscuous biotin ligase fusion protein identifies proximal and interacting proteins in mammalian cells. J Cell Biol. 2012;196(6):801–10.

